# Centiloid Calibration and Application of MUSE-Based Amyloid PET Quantification for [¹¹C]PiB and [¹⁸F]Florbetapir (AV-45)

**DOI:** 10.64898/2026.11.23.26367751

**Authors:** Saba Ishrat, Hoang Nam Le, Bahram Yaghooti, Ram Sapkota, J. Philip Miller, Deepa S. Thakuri, Murat Bilgel, Susan Resnick, Arman Rahmim, Christos Davatzikos, Dean F. Wong, Ganesh B. Chand, the Alzheimer’s Disease Neuroimaging Initiative

**Author notes:** These authors contributed equally to this work (Saba Ishrat, Hoang Nam Le). Data used in preparation of this article was obtained from the Alzheimer’s Disease Neuroimaging Initiative (ADNI) database (adni.loni.usc.edu). As such, the investigators within the ADNI contributed to the design and implementation of ADNI and/or provided data but did not participate in the analysis or writing of this report. A complete listing of ADNI investigators can be found at: http://adni.loni.usc.edu/wp-content/uploads/how_to_apply/ADNI_Acknowledgement_List.pdf. Corresponding authors: Saba Ishrat, DPhil, Hoang Nam Le, PhD, and Ganesh B. Chand, PhD.

## Abstract

**Introduction:** Amyloid positron emission tomography (PET) quantifies cerebral β-amyloid in Alzheimer’s disease (AD), but tracer- and processing-dependent variability limits comparability. We evaluated Centiloid transformations for multi-atlas segmentation (MUSE)-based [¹¹C]Pittsburgh compound B (PiB) and [¹⁸F]florbetapir (AV-45) PET.

**Methods:** Standardized uptake value ratio (SUVr) Centiloid transformations without partial-volume correction (NoPVC) and with parallel level set regularization-based PVC (PLS-PVC) were calibrated using the original Klunk Centiloid PiB reference dataset (N = 79) and paired PiB– AV-45 Navitsky dataset (N = 43). Centiloid transformations were evaluated in Alzheimer’s Disease Neuroimaging Initiative (ADNI; AV-45) and secondarily in Knight Alzheimer Disease Research Center (Knight-ADRC; PiB) cohorts. Outcomes included AD–cognitively normal (CN) separation and clinical associations. Scanner effects were examined in ADNI.

**Results:** MUSE-derived PiB SUVr measurements corresponded strongly with reference measurements (R² = 0.963, ICC(A,1) = 0.981), and PiB–AV-45 cross-tracer correspondence was high (R² = 0.873–0.884). NoPVC separated AD from CN in Knight-ADRC PiB (Hedges’ *g* = 1.907) and ADNI AV-45 (*g* = 0.892). PLS-PVC yielded similar separation in Knight-ADRC (Δ*g* = −0.027; 95% CI −0.122 to 0.023) and modestly greater separation in ADNI (Δ*g* = 0.053; 95% CI 0.038– 0.068). Scanner-associated variance represented 33.5% of total Centiloid variance in ADNI; on scanners other than one older model, AD–CN separation was larger (*g* = 1.31). Both approaches showed clinical associations.

**Discussion:** MUSE Centiloid transformations provide a framework for expressing PiB and AV-45 measurements in Centiloid units across independent cohorts. Scanner and processing differences warrant scanner-specific quality checks and cautious interpretation.

## 1. Introduction

Amyloid-β plaque deposition is a hallmark neuropathological feature of Alzheimer’s disease (AD) and related dementia, and amyloid positron emission tomography (PET) enables its detection and quantification *in vivo*.^1^ Centiloid-standardized amyloid PET measurements are increasingly used in research and clinical trials, including for patient selection, confirmation of amyloid pathology, and monitoring of amyloid-targeting therapies.^2^ Radiotracers include [¹¹C]Pittsburgh compound B (PiB) and the longer-lived ¹⁸F-labeled florbetapir (AV-45).^1,3–6^ However, quantitative measurements are not directly interchangeable across tracers and studies because of differences in tracer characteristics, acquisition protocols, scanner differences, anatomical definitions, and image-processing methods.^7,8^

To improve comparability, the original Centiloid framework standardizes amyloid PET measurements onto a common scale, with mean amyloid burden in young cognitively normal (CN) individuals defined as 0 Centiloid (CL) and that in typical patients with AD defined as 100 CL.^9^ The framework was originally established using PiB with a global cortical target and whole-cerebellum reference region and has subsequently been extended to other tracers and analytical approaches.^9–12^ Because differences in image processing and anatomical definitions can introduce systematic measurement differences, method-specific transformations are required. The MUlti-atlas region Segmentation utilizing Ensembles (MUSE) provides automated anatomical parcellation of structural MRI.^13–16^ MUSE outperforms segmentation techniques that use a single atlas, particularly in multi-site settings, across age ranges, and when segmenting both cortical and subcortical regions.^17^ MUSE has thus been increasingly applied in multi-cohort consortium studies of brain diseases,^18–22^ including analyses of brain aging,^23^ imaging-genetic subtyping,^24^ prediction of non-AD co-pathologies,^25^ neuropathology-confirmed imaging signatures of dementia,^26^ and cerebral small vessel disease,^27^ among others. However, a MUSE framework for expressing amyloid PET in CL units has not been established.

PET quantification is affected by partial-volume effects arising from limited spatial resolution, which can result in spill-in and spill-out of tracer signal across tissue boundaries and may be particularly relevant in the presence of cerebral atrophy.^28,29^ Partial-volume correction (PVC) is intended to reduce these effects, although its impact depends on the correction method and accuracy of the underlying image registration and segmentation.^29–34^ Centiloid values with and without PVC have been reported for the FreeSurfer-based PUP pipeline.^35,36^ We therefore hypothesized that PVC would improve AD–CN group separation by reducing signal dilution. To test this within the MUSE framework, we compared processing without partial-volume correction (NoPVC) with parallel level set regularization-based partial-volume correction (PLS-PVC)^37^ with respect to AD–CN separation and associations with cognitive and clinical severity. PLS-PVC was selected because it outperforms several other evaluated PVC methods.^37^ Because PVC was not incorporated into the original Centiloid standard, separate transformations were established for NoPVC and PLS-PVC measurements.

Cross-tracer harmonization presents an additional challenge. Quantitative approaches have been developed to standardize AV-45 standardized uptake value ratio (SUVr) measurements,^38^ and PiB and AV-45 measurements are strongly associated, although their quantitative relationships depend on tracer characteristics, imaging, and processing methodology.^7^ Navitsky et al. made paired PiB and AV-45 scans publicly available that enable AV-45 measurements to be expressed in CL units.^39^ This can be achieved by extending MUSE quantification for PiB to AV-45, which requires a cross-tracer conversion.

The study objectives were to (1) establish PiB-to-Centiloid transformations for our MUSE-based PET processing pipeline using the original Centiloid reference dataset^9^; (2) extend the PET Centiloid framework for MUSE to AV-45 through cross-tracer calibration using paired PiB and AV-45 data from the Navitsky dataset; (3) evaluate PiB transformations in an independent Knight Alzheimer Disease Research Center (Knight-ADRC) cohort by examining AD–CN separation and associations with cognitive and clinical severity scores; and (4) evaluate AV-45 transformations in an independent, highly diverse multi-scanner Alzheimer’s Disease Neuroimaging Initiative (ADNI) cohort, including scanner effects. Separate transformations were derived for NoPVC and PLS-PVC measurements, with NoPVC as the primary analysis and PLS-PVC as a secondary exploratory analysis. Additionally, we derived MUSE-based PiB and AV-45 Centiloid transformations for six predefined regional gray matter (GM) targets and individual GM ROIs. We hypothesized that MUSE-based PET Centiloid measures would show strong correspondence with published reference Centiloid measurements and demonstrate expected diagnostic and clinical associations in independent cohorts.

## 2. Materials and Methods

### 2.1 Standardized Calibration and Application Datasets

#### 2.1.1 Calibration datasets

The original Centiloid calibration dataset described by Klunk et al.^9^ was used for PiB calibration. The dataset included 34 young CN participants (YC-0) and 45 participants with AD (AD-100), defining the 0- and 100 anchors for the original Centiloid scale. The standard Centiloid method used PiB SUVr measured from 50–70 minutes after injection with a global cortical target and whole-cerebellum reference region defined using standardized volumes of interest (VOIs) in MNI152 space with SPM8, and SUVr was calculated with the GAAIN cortical VOI, defined from voxels with greater uptake in AD than in young controls, and a whole-cerebellum VOI.^9^ In contrast, the present study used MUSE-derived anatomical parcellations, with a global cortical target and cerebellar GM reference region, for PET quantification. The same reference dataset was downloaded from the Global Alzheimer’s Association Interactive Network (GAAIN; http://www.gaain.org) and processed at the Neuroimaging Informatics and Artificial Intelligence (NeuroAI) Laboratory, Washington University School of Medicine, using the MUSE-based PET quantification framework^14–16^ to establish MUSE PiB-to-Centiloid transformations. Rather than regressing MUSE SUVr on standard-method SUVr (Level-2 procedure^9^), CL values were obtained by direct scaling to the MUSE-derived YC-0 and AD-100 means.

Cross-tracer calibration used the paired PiB and AV-45 dataset described by Navitsky et al.^39^ downloaded from the GAAIN (http://www.gaain.org) and processed as outlined above. In that study, both the standard Centiloid approach, using cortical target and whole-cerebellum reference VOIs in MNI152 space, and an alternative florbetapir-specific PET-template-based approach using Avid-defined VOIs were evaluated.^38,39^ Paired PiB and AV-45 scans were originally acquired in 47 participants spanning a broad range of amyloid burden; one participant was excluded from the published analysis because of an AV-45 acquisition error, leaving 46 participants. PiB and AV-45 PET scans were acquired 50–70 and 50–60 minutes after injection, respectively, on separate days, with a mean interscan interval of 18 ± 20 days. For the present MUSE analysis, three additional participants were excluded because of incomplete acquisition information, resulting in a final paired sample of 43 participants. Both PiB and AV-45 scans were processed using the MUSE-based PET quantification framework^14,15^ to obtain NoPVC and PLS-PVC measurements.

#### 2.1.2 Knight-ADRC application dataset

The Knight-ADRC at Washington University in St. Louis maintains a longitudinal registry of community-dwelling volunteers spanning CN older adults, individuals with mild cognitive impairment, and individuals with symptomatic AD dementia.^40^ Established in 1985, the Knight-ADRC is an NIH-funded Alzheimer’s Disease Research Center. Of the 39 participants with 30-min dynamic PiB PET acquired 30–60 min after injection, 36 had a diagnosis of AD dementia or CN at a clinical assessment within 180 days of PET; amyloid status was not used for group assignment. Three of these were excluded after quality control: one because global SUVr could not be quantified and two because of large changes in global SUVr between uptake-window definitions that suggested registration or excessive motion errors. The analysis cohort comprised 33 participants (18 AD dementia, 15 CN). AD participants were older than CN participants (72.44 ± 5.43 vs 66.00 ± 6.98 years, *P* = 0.007), whereas education (15.39 ± 2.68 vs 16.27 ± 2.49 years, *P* = 0.338) and sex (44.4% vs 46.7% female, *P* = 1.000) did not differ. Age, sex, and education were included as covariates in the adjusted clinical analyses. Scans were acquired on three Siemens systems (Biograph mMR, n = 24; Biograph Vision 600 Edge, n = 6; Biograph 40 TruePoint, n = 3). T1-weighted MRI was acquired at 3T. Acquisition details are provided in **Supplementary Table 1**.

#### 2.1.3 ADNI application dataset

A subset of ADNI AV-45 PET was analyzed to evaluate CL conversions. The considered cohort (N = 238; 119 AD, 119 CN) was balanced for age (73.66 ± 8.08 vs 73.8 ± 6.25 years, *P* = 0.882), education (15.82 ± 2.76 vs 16.28 ± 2.44 years, *P* = 0.172), and sex (43.7% vs 48.7% female, *P* = 0.516). AD and CN groups were defined by ADNI clinical diagnosis; amyloid status was not used for group assignment. T1-weighted MRI was acquired at 3T (**Supplementary Table 1**). Scans were acquired on seven scanner families (see **Scanner effects in multisite analysis**).

Data used in the preparation of this article were obtained from the ADNI database (*adni.loni.usc.edu*). The ADNI was launched in 2003 as a public-private partnership, led by Principal Investigator Michael W. Weiner, MD. The original goal of ADNI was to test whether serial MRI, PET, other biological markers, and clinical and neuropsychological assessment can be combined to measure the progression of mild cognitive impairment (MCI) and early AD. The current goals include validating biomarkers for clinical trials, improving the generalizability of ADNI data by increasing diversity in the participant cohort, and providing data concerning the diagnosis and progression of Alzheimer’s disease to the scientific community. For up-to-date information, see *adni.loni.usc.edu*.

### 2.2 The MUSE-based amyloid PET processing framework

#### 2.2.1 MRI processing and MUSE segmentation

T1-weighted MR images were processed with MUSE version 3.1.0.^13^ Images were reoriented and corrected for intensity non-uniformity with N4, skull-stripped using MUSE with brain-extraction templates, and further bias-corrected with FSL FAST.^41,42^ Anatomical labels were then generated with MUSE, a multi-atlas method that registers the most similar atlases from a set of 35 manually labeled reference brains to each participant using two deformable registration algorithms DRAMMS and ANTs, and fuses the resulting candidate labels with intensity- and similarity-weighted label fusion. Default MUSE parcellation settings were used, except that both registration algorithms were applied. The resulting labels defined the global cortical target, the regional GM targets, the individual GM ROIs and the reference region. Full processing parameters are provided in **Supplementary Methods A**.

#### 2.2.2 Amyloid PET processing and global cortical SUVr

Quality checks, including assessment of motion, image artifacts, anterior-posterior commissure alignments, and field-of-view coverage, were applied to all PET and MRI scans.

For the Centiloid PiB calibration dataset, PET data acquired 50–70 min post-injection were used; for the paired Navitsky dataset, PiB and AV-45 data acquired 50–70 and 50–60 min post-injection were used, and for the application cohorts, data acquired 50–60 min (Knight-ADRC PiB) and 50–70 min (ADNI AV-45) post-injection were used. Dynamic PET frames were realigned with SPM12 and averaged over the uptake window, weighted by frame duration. Frames were included when their start and end times fell within the uptake window, allowing a tolerance of 0.5 min at the window boundaries to accommodate small differences in recorded frame timing. For the Knight-ADRC PiB cohort, which was acquired 30–60 min after injection, this corresponded to the last 10 min of the acquisition; for ADNI AV-45, acquired 50–70 min after injection as four 5-min frames, all frames were averaged. The averaged window was verified for each participant from the frame timing information. Injected dose, frame binning, and reconstruction for each application cohort are summarized by scanner in **Supplementary Table 1**. Reconstructed dynamic frames were used as provided by each site, without additional processing other than the frame realignment described above. The mean PET image was rigidly coregistered to the skull-stripped T1-weighted MRI using FSL FLIRT with mutual information,^43^ and the MUSE labels were transferred to each participant’s native PET space, where all regional measurements were obtained.

Amyloid burden was quantified as global cortical SUVr, using the MUSE mean cortical composite (36 cortical regions per hemisphere; **Supplementary Methods B**) as the target and cerebellar GM (MUSE cerebellar cortex, excluding the vermis and cerebellar white matter) as the reference region.^4,15,16^

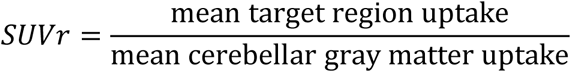

Each PET scan was processed with two implementations of the pipeline. NoPVC was designated the primary analysis because partial-volume correction is not part of the original Centiloid method; PLS-PVC transformations are provided for users of PVC-processed data and are reported as a secondary exploratory analysis. For NoPVC, regional uptake was quantified without partial-volume correction. For PLS-PVC, partial-volume effects were corrected with a deconvolution method using parallel level set regularization, which incorporates anatomical information from the T1-weighted MRI,^37^ as previously applied within our MUSE amyloid PET framework.^14,44^ PLS-PVC used an isotropic point-spread function of 4.4 mm full width at half maximum, a regularization weight of 48 and 100 iterations, for all datasets.

In addition to the global cortical measurement, regional SUVr measurements were obtained for six predefined MUSE GM targets: posterior cingulate, lateral temporal GM, frontal GM, precuneus, parietal GM, and occipital GM. These regional targets were selected based on their broad correspondence with regions evaluated in prior amyloid PET studies, although the anatomical definitions of the MUSE targets may differ from those used in previous studies.^45–47^ The anatomical composition of these predefined regional targets is provided in **Supplementary Table 2**. Regional SUVr measurements were calculated using the same PET-processing framework. Centiloid transformations were derived for these six predefined regional GM targets separately for NoPVC and PLS-PVC. These regional values represent regional measurements expressed on a Centiloid-referenced scale and should not be interpreted as equivalent to the standard global cortical Centiloid measurement. The regional analysis was further extended to individual MUSE GM ROIs.

### 2.3 PiB–AV-45 cross-tracer conversion

The cross-tracer calibration procedure was based on the general cross-tracer framework described by Klunk et al. and subsequently implemented for AV-45 by Navitsky et al.^9,39^ In the calibration datasets, global cortical SUVr values for PiB and AV-45 were calculated using the MUSE framework for NoPVC (primary) and PLS-PVC (secondary/exploratory). The calibration procedure was performed separately for NoPVC and PLS-PVC.

First, the PiB-to-Centiloid anchors for 0 CL and 100 CL were established using the original Centiloid reference dataset comprising 34 young cognitively normal participants (YC-0) and 45 participants with AD (AD-100),

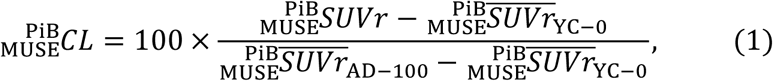

where 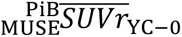 and 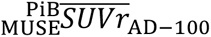 represent the corresponding means of SUVr values estimated using MUSE segmentations for the 34 young cognitively normal participants and 45 participants with AD, respectively. In the paired Navitsky dataset (N = 43), AV-45 SUVr was regressed on PiB SUVr,

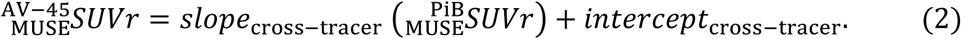

The fitted relationship was inverted to obtain the PiB-equivalent SUVr corresponding to each AV-45 measurement

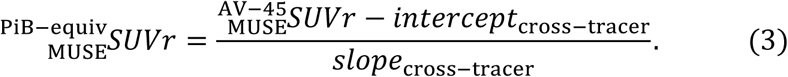

The resulting PiB-equivalent SUVr was then substituted into the fixed MUSE PiB-to-Centiloid transformation in **Equation 1**, resulting in a CL transformation for AV-45:

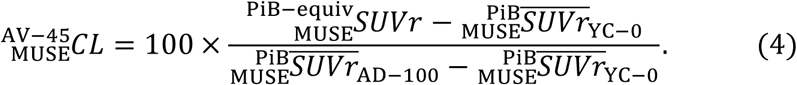

The paired Navitsky dataset therefore provided the cross-tracer relationship required to transform AV-45 measurements onto the same Centiloid scale as that of PiB within the MUSE-based PET quantification framework. The same calibration procedure was repeated for each predefined regional GM target and individual GM ROI.

### 2.4 Scanner effects in multisite analysis

The ADNI AV-45 cohort was acquired on several PET systems. We therefore examined whether scanner contributed to variation in Centiloid values. Participants were grouped into seven scanner families based on the PET manufacturer and model recorded by ADNI: GE Advance, GE Discovery, Philips, Siemens ECAT HR+, Siemens 1094, Siemens HI-REZ, and Siemens Biograph64/mCT. Within each scanner family, we summarized Centiloid values by diagnostic group. We also report the AD–CN difference and Hedges’ g for each family. Because the HR+ showed a distinctly smaller AD–CN difference, the same summaries were repeated comparing HR+ with all other scanners combined.

To quantify the scanner effect, we fitted an ordinary least-squares model of Centiloid on scanner family, diagnosis, age, and sex. The scanner share of variance was defined as the type II sum of squares for scanner family divided by the total sum of squares of Centiloid. To test whether group separation differed across scanners, we added a diagnosis × scanner interaction term and assessed it with a type II F-test. Both analyses were repeated with HR+ vs other scanners as the grouping variable.

As a sensitivity analysis, scanner effects were removed with ComBat^48,49^ (neuroCombat v0.2.12). The primary harmonized version and two further variants are described in **Supplementary Methods C**.

For each version, AD and CN means and SDs, Hedges’ g, AUC, and the residual scanner share of variance were reported. Scanner-effect analyses were performed for NoPVC and repeated for PLS-PVC Centiloid values; the harmonization sensitivity analysis was performed on NoPVC Centiloid values only. Harmonized values were used only in this sensitivity analysis. All primary results are reported on the unharmonized Centiloid scale.

### 2.5 Scope of the transformations and implementation criteria

Adopters of MUSE Centiloid should follow the three-level structure of The Centiloid Project^9^ when they want to express their data in MUSE CL units using the transformations derived by this work. The transformations given by the MUSE CL equations apply only to data processed with the same method: MUSE segmentation of T1-weighted MRI, the MUSE mean cortical target, cerebellar GM reference, and NoPVC or PLS-PVC processing. They were calibrated with PiB acquired 50–70 min and AV-45 acquired 50–60 min after injection.^9,39^ Applying them to other uptake windows, as in our application cohorts, assumes that the SUVr–CL relationship does not depend on the window.^50^ Under these conditions, a site can use the transformations as a Centiloid Level-3 analysis (exact reproduction of a calibrated method) without new calibration data. Before applying them, a site should verify that its pipeline reproduces our values for each tracer it intends to use. For PiB, the site processes the 34 YC-0 and 45 AD-100 scans of the original Centiloid dataset, and the mean SUVr of each group should fall within 2% of our reported values. For AV-45, the site processes the AV-45 scans of the 43 Navitsky participants and compares the resulting measurements with the corresponding MUSE-derived reference values. For both tracers, the site’s CL values are compared with ours, with a slope of 0.98–1.02, an intercept within ±2 CL, and R² > 0.98. Once the pipeline is verified, global cortical SUVr is converted to CL without further fitting.

Any departure from this method requires a new Level-2 calibration. This includes a different PVC algorithm, target or reference region, segmentation version, acquisition window, or tracer. Level-2 calibration requires at least 25 participants with PiB (50–70 min) and the new tracer or method acquired within 3 months of each other following the guideline established by Klunk et al.^9^ The site then reports R² (expected > 0.70), the slope and intercept, and the relative variance in young controls compared with PiB. The regional transformations apply only to the corresponding MUSE region and processing. Each was anchored to that region’s own YC-0 and AD-100 means, so regional values are not equivalent to global CL.

### 2.6 Statistical analysis

Agreement between MUSE-based CL and published CL values was assessed with correlation analysis and the paired difference (MUSE − published), summarized as mean ± SD with 95% limits of agreement (mean ± 1.96 SD), and with the intraclass correlation coefficient for absolute agreement (ICC(A,1); two-way random effects, single measures). Group differences in age, years of education, and Mini-Mental State Examination (MMSE) were tested with Welch’s t-test, whereas group differences in Clinical Dementia Rating (CDR) global score and CDR Sum of Boxes (CDR-SB) were evaluated using the Mann–Whitney U test. Cross-tracer correspondence was summarized using Pearson correlation coefficients (*r*) and the coefficient of determination (R²).

Associations between CL values and cognitive and clinical severity scores were evaluated using Spearman’s rank correlations (*ρ*) within the independent Knight-ADRC PiB and ADNI AV-45 cohorts. Cognitive and clinical measures included the CDR global score, CDR-SB, and MMSE. Clinical assessments obtained within 180 days of PET were included. Partial Spearman’s correlations were additionally calculated after adjustment for age, sex, and education. For the regional application analyses, adjusted associations with CDR-SB and MMSE were evaluated separately for each of the six predefined regional GM targets.

Diagnostic separation between participants with AD dementia and CN participants was quantified using unadjusted bias-corrected Hedges’ *g*, and group differences were evaluated using Welch’s independent-samples *t* tests. For the regional analyses, diagnostic group comparisons were performed separately for each of the six predefined regional GM targets. Multiple-comparison correction was applied across the regional tests using the Benjamini– Hochberg false discovery rate (FDR) procedure, and adjusted significance values are reported as q values.

Because NoPVC and PLS-PVC measurements were obtained from the same participants, between-method differences in unadjusted and adjusted Spearman’s correlation coefficients and Hedges’ *g* were evaluated using paired subject-level bootstrap resampling with 10,000 repetitions. Differences were calculated as PLS-PVC minus NoPVC, and percentile 95% confidence intervals were obtained from the bootstrap distributions.

## 3. Results

### 3.1 MUSE-derived PiB Centiloid and cross-tracer calibration

In the original Centiloid reference dataset (N = 79; 34 YC-0 and 45 AD-100 participants), MUSE-derived PiB SUVr measurements showed strong correspondence (*r* = 0.981; R² = 0.963) when compared with published standard-method SUVr values (**Table 1**, **Figure 1A**). For the corresponding CL measurements, absolute agreement was high (ICC(A,1) = 0.981, 95% CI 0.971–0.988), with a mean difference of 0.0 ± 10.2 CL. Mean NoPVC PiB SUVr values for the YC-0 and AD-100 groups were 1.084 and 1.953 (**Table 1**). The resulting MUSE PiB-to-Centiloid transformations were:

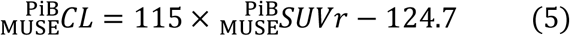

**Figure 1.**
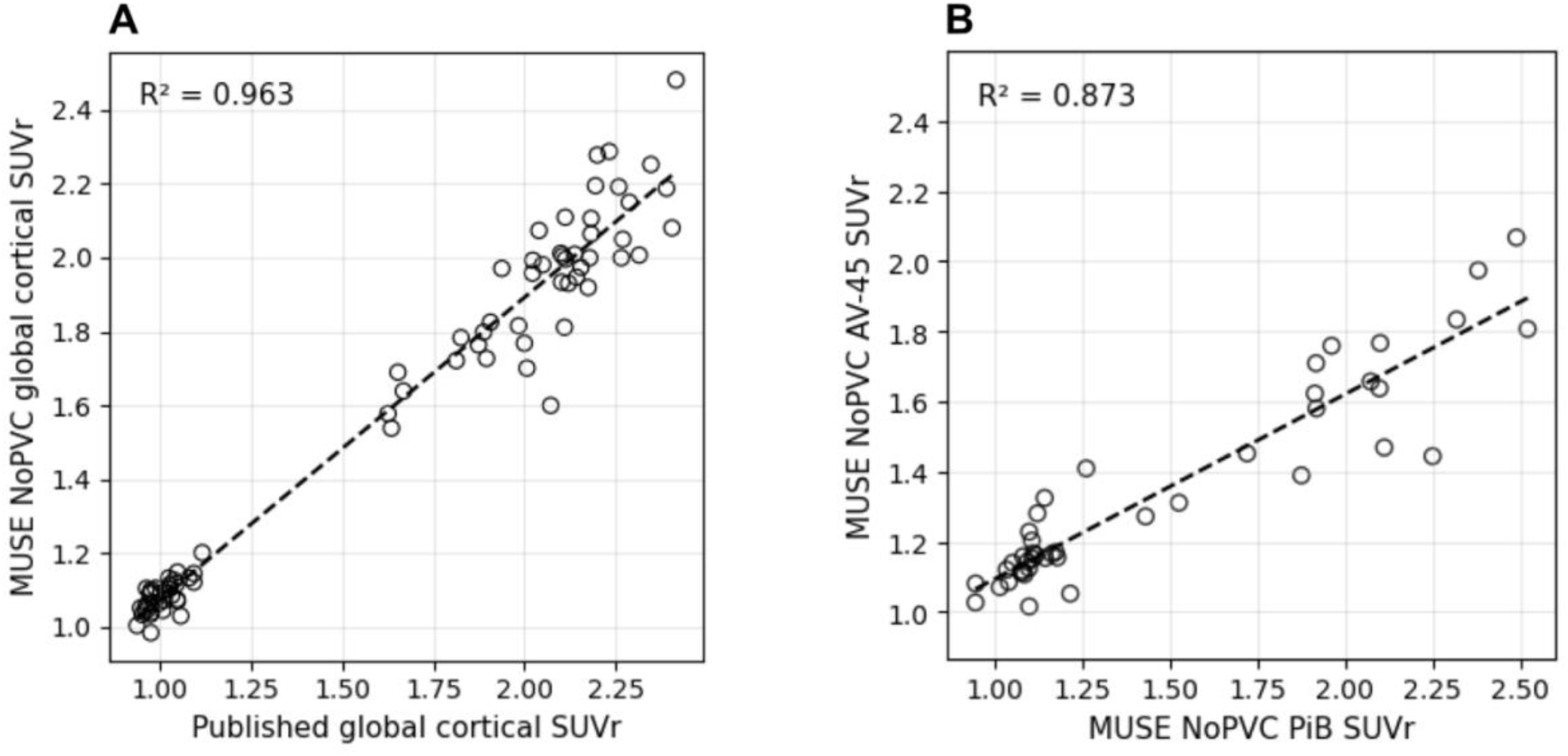
(A) Correspondence between published and MUSE-derived PiB SUVr measurements in the original Centiloid (CL) reference dataset (N = 79; *r* = 0.981; R² *=* 0.963; slope = 0.818; intercept = 0.257). The scatterplot compares published global cortical SUVr measurements obtained using the whole-cerebellum reference region with MUSE NoPVC measurements obtained using the cerebellar GM reference region. (B) PiB–AV-45 cross-tracer relationship in the paired Navitsky dataset (N = 43, *r* = 0.935; R² *=* 0.873; slope = 0.527; intercept = 0.566), using MUSE-derived SUVr measurements without PVC. Dashed lines represent fitted linear regressions.

**Table 1.** MUSE-based PiB Centiloid and PiB–AV-45 cross-tracer calibration.

|  | Published | NoPVC | PLS-PVC |
|---|---|---|---|
| **PiB Centiloid calibration (N = 79)** |  |  |  |
| YC-0 SUVr, mean (SD) | 1.010 (0.046) | 1.084 (0.045) | 1.073 (0.045) |
| AD-100 SUVr, mean (SD) | 2.076 (0.201) | 1.953 (0.206) | 2.032 (0.232) |
| YC-0 CL, mean (SD) | 0 (4.336) | 0 (5.173) | 0 (4.688) |
| AD-100 CL, mean (SD) | 100 (18.877) | 100 (23.708) | 100 (24.187) |
| Reference correspondence, *r* | — | 0.981 | — |
| Reference correspondence, R² | — | 0.963 | — |
| CL agreement, slope/intercept | — | 1.0/−0.2 | — |
| Mean CL difference ± SD | — | 0.0 ± 10.2 (*P* = 0.997) | — |
| ICC(A,1) (95% CI) | — | 0.981 (0.971–0.988) | — |
| **PiB–AV-45 cross-tracer calibration (N = 43)** |  |  |  |
| Cross-tracer regression slope | 0.536 | 0.527 | 0.524 |
| Cross-tracer regression intercept | 0.502 | 0.566 | 0.555 |
| Cross-tracer *r* | 0.946 | 0.935 | 0.940 |
| Cross-tracer R² | 0.894 | 0.873 | 0.884 |

Values are presented as mean (SD) unless otherwise indicated. Published Level-1 calibration values correspond to the standard Centiloid processing described by Klunk et al. PiB calibration was performed using the original Centiloid reference dataset (N = 79; 34 YC-0 and 45 AD-100 participants), and PiB–AV-45 cross-tracer calibration was performed using the paired Navitsky dataset (N = 43). Separate transformations were derived for NoPVC and PLS-PVC measurements. Reference correspondence represents the association between MUSE-derived and reference measurements across the original Centiloid dataset. Mean CL difference represents paired difference between MUSE-derived CLs and published values, MUSE – published. Cross-tracer regression represents linear regression of MUSE-derived AV-45 SUVr on MUSE-derived PiB SUVr in participants with complete paired scans. Abbreviations: AD-100, Alzheimer disease group defining the 100-Centiloid anchor; AV-45, [<sup>18</sup>F]florbetapir; CL, Centiloid; MUSE, MUlti-atlas region Segmentation utilizing Ensembles; PiB, [<sup>11</sup>C]Pittsburgh compound B; PLS-PVC, parallel level set partial-volume correction; SUVr, standardized uptake value ratio; NoPVC, without partial-volume correction; YC-0, young cognitively normal group defining the 0-Centiloid anchor; ICC, intraclass correlation coefficient.

The corresponding PLS-PVC transformation was CL = 104.3 × SUVr − 111.9. The NoPVC transformations are illustrated in **Figure 2** and were used for subsequent AV-45 cross-tracer conversion.

**Figure 2.**
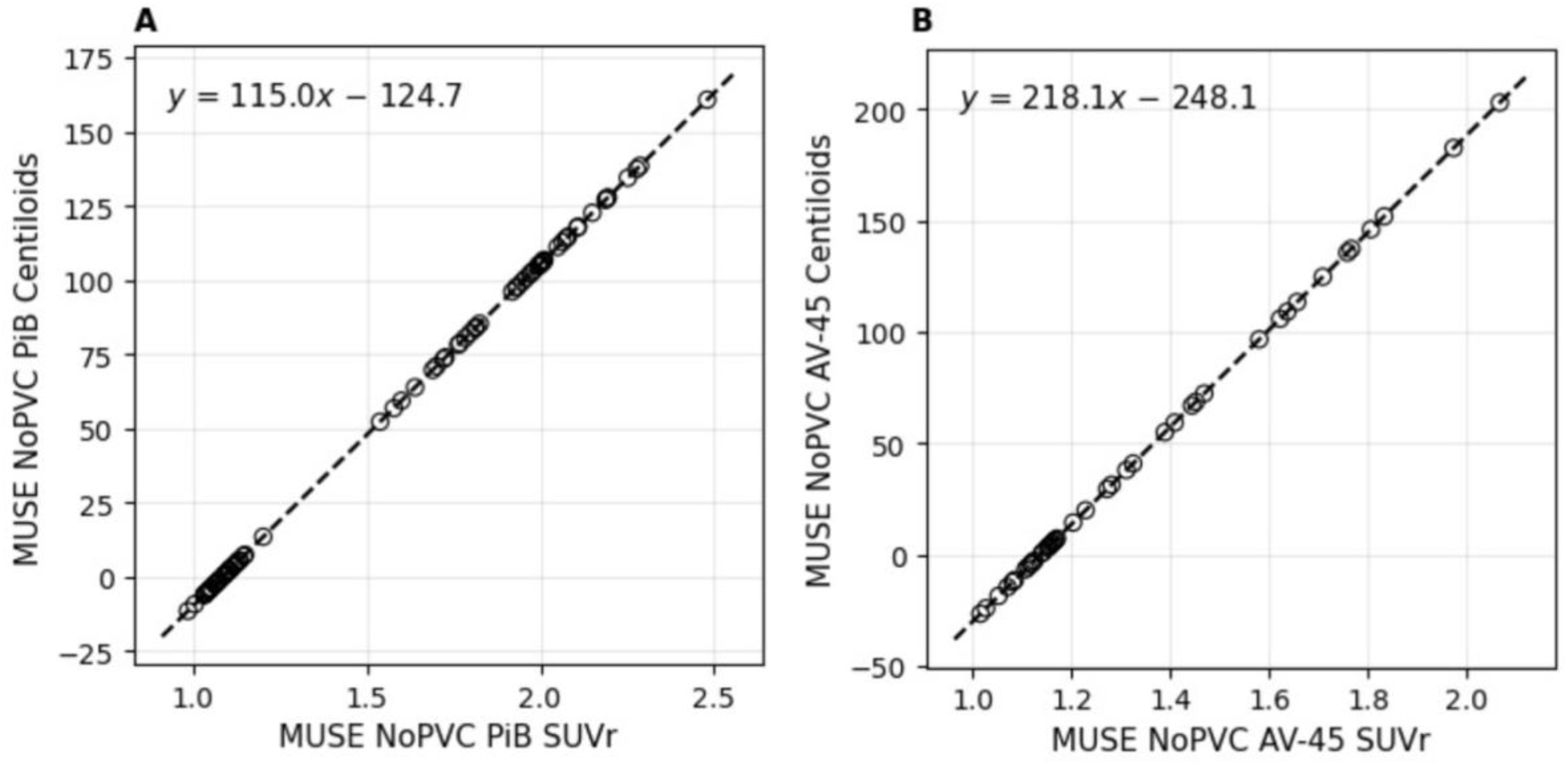
MUSE PiB and AV-45 SUVr-to-Centiloid transformations for NoPVC processing. Scatterplots illustrate the relationships between MUSE-derived global cortical SUVr measurements and the corresponding calculated Centiloid (CL) values for (A) PiB, established using the original Centiloid reference dataset (N = 79); (B) AV-45, derived through cross-tracer calibration using the paired Navitsky dataset (N = 43). Open circles represent individual participant measurements, and dashed lines represent the corresponding SUVr-to-Centiloid transformations. The equations are displayed within each panel.

In the paired Navitsky dataset, MUSE-derived AV-45 SUVr showed strong correspondence with MUSE-derived PiB SUVr for both processing approaches. Linear regression of AV-45 SUVr on PiB SUVr yielded a slope and intercept of 0.527 and 0.566 for NoPVC (*r* = 0.935; R² = 0.873), as well as 0.524 and 0.555 for PLS-PVC (*r* = 0.940; R² = 0.884) (**Table 1**; **Figure 1B**). **Equation 6** gives the resulting AV-45-to-Centiloid transformation for NoPVC; the corresponding PLS-PVC transformation was CL = 199.0 × SUVr − 222.5:

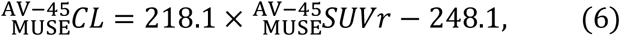

In the original Centiloid reference dataset, MUSE-derived NoPVC CL values agreed closely with the published CL values (slope = 1.0, intercept = −0.2 CL, R² = 0.963; **Figure 3A**). Because both scales were anchored to the same YC-0 and AD-100 participants, the near-unity slope and near-zero intercept are largely expected by construction; the high R², however, indicates close participant-level agreement despite the different target and reference regions. In the independent paired Navitsky dataset, MUSE-derived CL values remained highly correlated with the published values but were systematically scaled. Slopes were 1.20 for PiB and 1.25 for AV-45 (R² = 0.971 and 0.974, respectively; **Figure 3B**). However, MUSE CL values for PiB and AV-45 were systematically higher than published values (mean difference 8.6–9.0 CL, SD 13.6–15.9 CL; *P* < 0.001), and absolute agreement was lower than the correlation (ICC(A,1) = 0.948–0.955, versus *r* = 0.986–0.987; **Supplementary Table 3**). Thus, although relative amyloid burden was preserved across methods, the absolute CL values depended on the underlying quantification approach. Participant-level MUSE-derived PiB SUVr and CL values for the calibration datasets are provided in **Supplementary Tables 4** and **5**.

**Figure 3.**
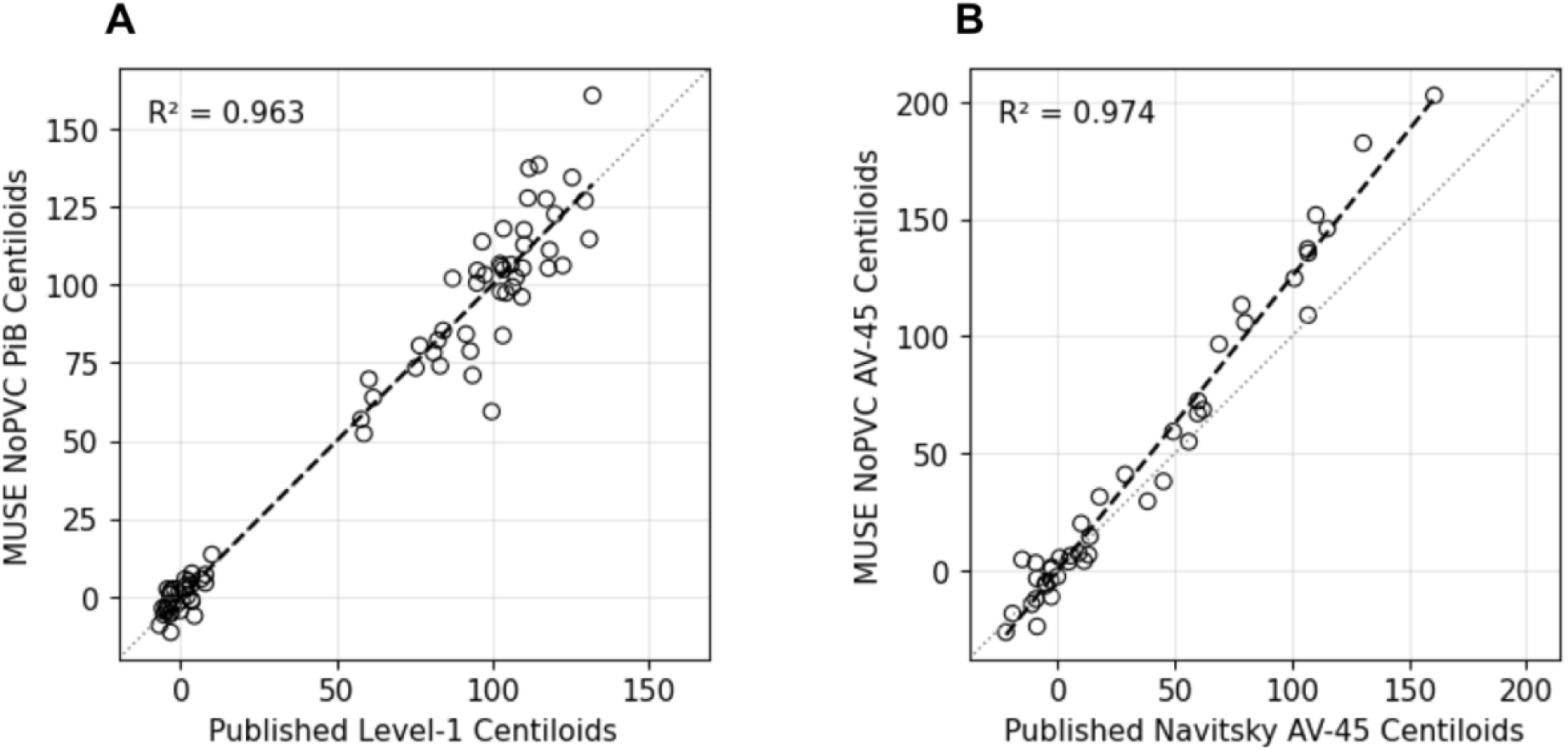
Correspondence of MUSE Centiloids (NoPVC) with published Centiloids for PiB and AV-45. **(A)** MUSE NoPVC PiB Centiloid versus the published Level-1 Centiloid values for the 79 participants of the original Centiloid reference dataset (*r* = 0.981; R² *=* 0.963; *P* = 5.38 × 10^-57^). MUSE Centiloids were derived from the MUSE global cortical SUVr (cerebellar GM reference) using the MUSE YC-0 and AD-100 anchors; published values use the standard Centiloid cortical ROI with a whole-cerebellum reference. **(B)** NoPVC MUSE AV-45 Centiloid versus the published AV-45 Centiloid values for 43 participants with paired PiB and florbetapir PET (*r* = 0.987; R² = 0.974; *P* = 4.26 × 10^-3^^4^). MUSE AV-45 Centiloids were calculated with the MUSE PiB anchors from (A), combined with the cross-tracer regression from the paired Navitsky data. Dashed lines show the least-squares regression.

### 3.2 Independent application of Centiloid transformations

#### 3.2.1 Knight-ADRC PiB application

Cohort characteristics of the application datasets are provided in **Supplementary Table 6**. The Knight-ADRC PiB application cohort comprised 33 participants, including 18 participants with AD dementia and 15 CN participants. NoPVC Centiloid values distinguished participants with AD dementia from CN participants (**Table 2**): mean CL values were 95.2 ± 66.5 and −2.6 ± 13.2, respectively, a difference of 97.8 CL (Hedges’ *g* = 1.907; Welch’s *P* = 8.04 × 10⁻⁶). PLS-PVC gave similar separation (*g* = 1.880; **Supplementary Table 7**), and the between-method difference was small, with a 95% confidence interval including zero (Δ*g* [PLS-PVC − NoPVC] = −0.027, 95% CI −0.122 to 0.023).

**Table 2.** Independent application of MUSE-derived Centiloid measurements (NoPVC) in the Knight-ADRC PiB and ADNI AV-45 cohorts.

|  | **Knight-ADRC**<br>**PiB** | **ADNI**<br>**AV-45** |
|---|---|---|
| AD dementia, CL, mean (SD) | 95.2 (66.5) | 63.9 (74.8) |
| CN, CL, mean (SD) | –2.6 (13.2) | 6.1 (52.6) |
| AD−CN mean difference, CL | 97.8 | 57.9 |
| Hedges’ *g* | 1.907 | 0.892 |
| Welch’s *P* | 8.04 × 10<sup>–6</sup> | 5.93 × 10<sup>–11</sup> |
| CDR-SB, adjusted *ρ* | 0.536 | 0.334 |
| Global CDR, adjusted *ρ* | 0.533 | 0.372 |
| MMSE, adjusted *ρ* | –0.392 | –0.364 |

Values are presented as mean (SD) unless otherwise indicated. Diagnostic separation was assessed between participants with AD dementia and CN participants (Knight-ADRC PiB, 18 AD and 15 CN; ADNI AV-45, 119 AD and 119 CN). Clinical associations are partial Spearman’s correlations adjusted for age, sex, and education. PLS-PVC results and between-method differences are provided in Supplementary Tables 7 and 8. Abbreviations: AD, Alzheimer disease; ADNI, Alzheimer’s Disease Neuroimaging Initiative; AV-45, [<sup>18</sup>F]florbetapir; CDR, Clinical Dementia Rating; CDR-SB, Clinical Dementia Rating Sum of Boxes; CN, cognitively normal; CL, Centiloid; MMSE, Mini-Mental State Examination; MUSE, MUlti-atlas region Segmentation utilizing Ensembles; PiB, [<sup>11</sup>C]Pittsburgh compound B; PLS-PVC, parallel level set regularization-based partial-volume correction; NoPVC, without partial-volume correction.

MUSE-derived PiB CL values were also associated with clinical and cognitive severity in the expected directions. After adjustment for age, sex, and education, NoPVC partial Spearman’s correlations were 0.536 for CDR-SB (*P* = 0.002), 0.533 for global CDR (*P* = 0.002), and −0.392 for MMSE (*P* = 0.035) (**Table 2**). Correlations with PLS-PVC were similar (0.512, 0.508, and −0.361; the MMSE association did not reach statistical significance, *P* = 0.055), and all PLS-PVC − NoPVC differences had 95% confidence intervals including zero (**Supplementary Table 7**).

#### 3.2.2 ADNI AV-45 application

The matched ADNI AV-45 application cohort comprised 238 participants, with 119 participants with AD dementia and 119 CN participants. NoPVC Centiloid separated the two groups (**Table 2**): mean CL values were 63.9 ± 74.8 in participants with AD dementia and 6.1 ± 52.6 in CN participants, a difference of 57.9 CL (Hedges’ *g* = 0.892; Welch’s *P* = 5.93 × 10⁻¹¹). The observed separation was weaker than in the Knight-ADRC PiB cohort, and CN values were considerably more dispersed; potential contributions from scanner-related variability are examined below, and other contributors are considered in the Discussion. PLS-PVC yielded modestly greater AD–CN separation (*g* = 0.944; Δ*g* [PLS-PVC − NoPVC] = 0.053, 95% CI 0.038 to 0.068; **Supplementary Table 8**).

Associations with clinical and cognitive measures were observed for both processing approaches. After adjustment for age, sex, and education, NoPVC partial Spearman’s correlations were 0.334 for CDR-SB, 0.372 for global CDR, and −0.364 for MMSE (all *P* < 10⁻⁶; **Table 2**). Correlations with PLS-PVC were slightly stronger (0.356, 0.394, and −0.387), with PLS-PVC − NoPVC differences of 0.022 (95% CI 0.013 to 0.032), 0.022 (95% CI 0.013 to 0.032), and −0.022 (95% CI −0.034 to −0.013), respectively (**Supplementary Table 8**).

NoPVC Centiloid values differed substantially across scanner families (**Supplementary Table 9**; **Supplementary Figure 1**). After adjustment for diagnosis, age, and sex, scanner family accounted for 33.5% of total Centiloid variance (type II scanner sum of squares divided by total sum of squares; *P* = 2.6 × 10⁻²³), and the AD–CN difference depended on scanner (diagnosis × scanner interaction, *P* = 1.3 × 10⁻⁴). This interaction was most pronounced for the Siemens ECAT HR+, on which both diagnostic groups had low Centiloid values with little separation: CN −34.5 ± 25.4 CL and AD −20.2 ± 28.3 CL, a difference of 14.3 CL (Hedges’ *g* = 0.53). The other scanner families combined showed a much larger separation, with CN 21.6 ± 52.1 CL and AD 96.2 ± 60.6 CL, a difference of 74.6 CL (*g* = 1.31). Within individual non-HR+ scanner families, Hedges’ *g* ranged from 1.26 to 2.06, closer to the separation observed in the Knight-ADRC PiB cohort (*g* = 1.907); the Siemens Biograph64/mCT estimate is based on only two CN participants. PLS-PVC Centiloid showed the same pattern (**Supplementary Table 9**). In a sensitivity analysis, ComBat harmonization reduced the scanner-associated share of total Centiloid variance from 33.5% to below 0.1%, and Hedges’ *g* increased from 0.892 to 1.116 (AUC 0.711 to 0.789), but separation on the HR+ was not restored (**Supplementary Methods C**; **Supplementary Table 11**). Scanner effects were not modeled in the Knight-ADRC PiB cohort, because only the Biograph mMR had at least two participants in both diagnostic groups; Centiloid values for this cohort are reported by scanner in **Supplementary Table 10**.

### 3.3 Regional MUSE Centiloid analyses

#### 3.3.1 Regional MUSE Centiloid and PiB–AV-45 cross-tracer calibration

The MUSE-based PET calibration framework was additionally extended from the global cortical measurement to six predefined regional GM targets: posterior cingulate, lateral temporal GM, frontal GM, precuneus, parietal GM, and occipital GM. Regional PiB and AV-45 Centiloid transformations were derived separately for NoPVC and PLS-PVC processing (NoPVC, **Table 3**; PLS-PVC, **Supplementary Table 12**). Each regional transformation was anchored to the corresponding regional YC-0 and AD-100 means, and regional CL values are not equivalent to global CL values. PiB–AV-45 cross-tracer correspondence varied across the six regional targets, with NoPVC R² ranging from 0.792 (posterior cingulate) to 0.875 (lateral temporal GM); PLS-PVC values were similar. Cross-tracer regression slopes and intercepts, and the PLS-PVC transformations, are provided in **Supplementary Table 12**. The analysis was further extended to individual MUSE GM ROIs, with the corresponding calibration and cross-tracer regression parameters provided in **Supplementary Table 13**. Cross-tracer correspondence varied substantially across individual ROIs; therefore, transformations for ROIs with low cross-tracer R² should be interpreted cautiously.

**Table 3.** MUSE-derived regional Centiloid transformations and PiB–AV-45 cross-tracer associations.

| **Gray matter target region** | **PiB CL equation** | **AV-45 CL equation** | **Cross-tracer R<sup>2</sup>** |
|---|---|---|---|
| Posterior cingulate | 113.7 × SUVr -140.5 | 225.2 × SUVr -279.4 | 0.792 |
| Lateral temporal GM | 119.4 × SUVr -123.3 | 225.4 × SUVr -244.9 | 0.875 |
| Frontal GM | 121.3 × SUVr -132.9 | 223.8 × SUVr -258.1 | 0.858 |
| Precuneus | 89.5 × SUVr -103.6 | 175.1 × SUVr -204.4 | 0.859 |
| Parietal GM | 117.9 × SUVr -129.9 | 224.6 × SUVr -259.3 | 0.853 |
| Occipital GM | 193.0 × SUVr -217.5 | 342.5 × SUVr -393.4 | 0.818 |

Regional Centiloid transformations are shown separately for NoPVC processing. Cross-tracer R² represents the coefficient of determination from the corresponding regional PiB–AV-45 regression. AV-45, [¹⁸F]florbetapir; CL, Centiloid; GM, gray matter; MUSE, MUlti-atlas region Segmentation utilizing Ensembles; PiB, [¹¹C]Pittsburgh compound B; PLS-PVC, parallel level set regularization-based partial-volume correction; R², coefficient of determination; SUVr, standardized uptake value ratio; NoPVC, without partial-volume correction.

#### 3.3.2 Regional application of MUSE-derived Centiloid measurements

The regional Centiloid transformations were subsequently applied to the independent Knight-ADRC PiB and ADNI AV-45 cohorts. In both cohorts, all six regional targets demonstrated significant AD–CN separation for both NoPVC and PLS-PVC measurements after multiple-comparison correction (**Supplementary Table 14**). After adjustment for age, sex, and education, regional Centiloid measurements were significantly associated with CDR-SB in all six regions in both cohorts and with MMSE in all six regions in ADNI after multiple-comparison correction; in Knight-ADRC PiB, the association with MMSE was significant in five of six regions, with occipital GM not reaching significance (*q* = 0.159 for NoPVC and 0.180 for PLS-PVC; **Supplementary Table 15**). The magnitude of diagnostic separation and clinical associations varied across anatomical regions and processing approaches.

## 4. Discussion

In this study, we established a MUSE-based framework for expressing PiB and AV-45 amyloid PET measurements in CL units across independent cohorts. Our MUSE PiB-to-Centiloid transformations were established using the original Centiloid reference dataset, and the framework was extended to AV-45 through cross-tracer calibration using paired PiB and AV-45 scans. MUSE-derived PiB measurements showed strong correspondence with the reference measurements, and strong relationships were observed between MUSE-derived PiB and AV-45 SUVr measurements. The resulting PiB and AV-45 transformations demonstrated expected AD–CN separation and associations with cognitive and clinical severity scores, although separation in the multi-scanner ADNI cohort was reduced by differences between PET scanners. Beyond the global cortical measurement, the framework was expanded to six predefined regional GM targets and individual GM ROIs. These findings support MUSE-based CL quantification while highlighting processing-dependent differences.

A central feature of the present framework is the calibration of MUSE-derived PET measurements to the Centiloid scale while retaining the anatomical definitions of MUSE. In the present study, the original Centiloid reference dataset comprising 34 YC-0 and 45 AD-100 participants was used to establish our MUSE PiB-to-Centiloid transformations. MUSE processing retained the MUSE-defined global cortical target and cerebellar GM reference regions. These transformations are specific to MUSE processing and are not interchangeable with other pipelines. Studies have similarly demonstrated that amyloid PET measurements obtained using different tracers and image-processing approaches can be related to the Centiloid scale through calibration procedures.^9–12^ The strong PiB–AV-45 correspondence obtained with MUSE, only slightly lower than that reported by Navitsky et al. for the standard Centiloid and Avid ROI approaches,^39^ suggests that strong cross-tracer correspondence can be maintained despite differences in target and reference region definitions.

Strong correspondence with published Centiloid values, however, does not imply equivalence of absolute CL values across processing methods. The scaling observed in the paired Navitsky dataset is particularly informative, because these participants were independent of the original PiB anchor dataset, although they were also used to derive the AV-45 cross-tracer transformation. Its presence indicates that the relationship between MUSE-based and standard measurements may not be fully constant across datasets, possibly reflecting scanner or image characteristics that affect the two region-definition approaches differently. Centiloid transformation therefore preserves the relative ordering of amyloid burden but does not make absolute values fully invariant to upstream processing.^51^ This matters when applying positivity thresholds or eligibility cutoffs from another pipeline or pooling CL values across pipelines.^2^ It also supports the replication step recommended by the Centiloid Project,^9^ which allows each site to verify its implementation before relying on absolute values.

Our PVC hypothesis was partly supported: PLS-PVC modestly improved AD–CN separation in ADNI but not Knight-ADRC PiB. In the Knight-ADRC PiB cohort, NoPVC and PLS-PVC measurements produced nearly identical AD–CN separation, and the paired between-method difference was small with a 95% confidence interval including zero. Partial-volume effects from limited PET resolution can produce spill-in and spill-out of signal across tissue boundaries, particularly in the presence of cerebral atrophy, and partial-volume correction may reduce these effects.^28–31^ At the same time, PVC may introduce sensitivity to segmentation errors and assumptions regarding regional signal distributions.^28^ In ADNI AV-45, PLS-PVC also yielded slightly stronger clinical associations than NoPVC. These findings do not establish a general PVC advantage but show that NoPVC and PLS-PVC can differ quantitatively even after CL transformation. NoPVC nevertheless remains relevant for methodological comparability with the original Centiloid framework.

The application cohorts showed how acquisition conditions affect calibration transfer.^51^ In the Knight-ADRC PiB cohort, AD–CN separation was large (*g* = 1.907). In the multi-scanner ADNI cohort, separation was relatively weaker (*g* = 0.892), and scanner family accounted for about one third of total CL variance. This effect was concentrated on the Siemens ECAT HR+, on which AD–CN separation was substantially reduced; within the other scanner families, separation (*g* = 1.26–2.06) was comparable to that in the Knight-ADRC cohort. The source of the HR+ effect could not be determined from the available data. Differences in spatial resolution, reconstruction, or attenuation and scatter correction that affect the MUSE cortical target and cerebellar GM reference differently are possible explanations.^52^ Scanner effects on Centiloid values have also been reported with the standard Centiloid pipeline. In participants imaged on two PET systems, a digital-detector scanner gave higher Centiloid values than a conventional scanner, and resolution harmonization corrected only about 20% of the difference.^53^ In florbetapir data, separation between amyloid-negative and amyloid-positive groups in Centiloid units increased with reconstructed spatial resolution.^54^ Harmonization methods such as ComBat can facilitate pooled multi-site analyses^55^ but cannot ensure absolute CL comparability across scanners. Approaches that address resolution directly, such as ADNI uniform-resolution images combined with a calibration performed at matching resolution, remain to be evaluated.^52^ Scanner-specific checks are therefore needed before MUSE Centiloid values are pooled across scanners.

In addition to scanner effects, group mean CL values in the application cohorts, particularly in ADNI, departed from the 0 and 100 anchors, and substantial variability was observed within diagnostic groups. This is partly expected because the anchors are defined by selected groups: young controls and participants with typical AD. The application cohorts were instead defined by clinical diagnosis. A substantial proportion of cognitively normal older adults have elevated cerebral amyloid. In an individual participant meta-analysis, amyloid positivity among cognitively normal individuals increased from approximately 10% at age 50 to 44% at age 90.^56^ This is consistent with a preclinical stage of AD that may begin many years before dementia.^57^ In ADNI, cognitively normal participants with elevated amyloid showed faster subsequent cognitive decline than those without.^58^ The inclusion of amyloid-positive cognitively normal individuals could therefore raise the CN mean and extend the upper tail of the CN distribution. Conversely, a minority of patients clinically diagnosed with AD dementia are amyloid-negative on PET.^59^ In ADNI, amyloid negativity was also supported by CSF Aβ42 measurements in most PET-negative AD participants with available CSF data, and these participants differed clinically and in other biomarkers from amyloid-positive patients, suggesting non-AD etiologies.^60^ Their inclusion could lower the AD mean and widen its distribution. Differences in Centiloid scaling may also contribute to variability expressed in CL units: because florbetapir has a smaller dynamic range, one SUVr unit corresponds to 218 CL for AV-45 compared with 115 CL for PiB in the MUSE NoPVC transformations, so equivalent absolute SUVr measurement errors would produce approximately 1.9-fold greater variation in CL units for AV-45.

The framework also extends global cortical quantification to anatomically defined regional measurements. Regional variation in PiB–AV-45 correspondence indicates that regional AV-45 transformations cannot inherit the global relationship. Regional transformations may be useful for characterizing early or spatially heterogeneous amyloid deposition that a global cortical composite may not capture. The occipital GM target showed the weakest AD–CN separation and was the only target without a significant association with MMSE in the Knight-ADRC PiB cohort. Amyloid deposition in the occipital cortex, particularly primary visual cortex, occurs later and is typically less pronounced in AD than in frontal and parietal association cortex,^61,62^ which may explain the weaker separation and the lack of association with MMSE. In addition, regional transformations for targets with a narrow YC-0 to AD-100 range, such as the occipital cortex, have steep slopes that amplify small between-cohort SUVr differences; this may contribute to the elevated occipital CL values in both diagnostic groups of the ADNI cohort, despite preserved AD–CN separation. Small regions are also more sensitive than the global composite to individual segmentation failures, so regional quality control is important. Because each regional transformation was anchored to that region’s own YC-0 and AD-100 means, regional CL values are not directly comparable with global CL.

The regional extension may also facilitate multimodal imaging analyses. Because MUSE provides a common anatomical parcellation that can support structural MRI measurements and regional PET quantification, the present framework may facilitate analyses of Centiloid-standardized regional amyloid burden alongside regional structural and other molecular imaging measures. Compared with conventional global cortical Centiloid approaches, the advantages of MUSE are anatomical rather than quantitative: native-space parcellation and regional and subcortical targets within the same framework used for structural MRI analyses.^23,25^

Several limitations should be acknowledged. First, the AV-45 cross-tracer calibration relied on 43 paired PiB–AV-45 participants, which may limit precision at the extremes of the amyloid range. Second, the application cohorts were defined by clinical diagnosis without amyloid confirmation, and the analyses evaluated diagnostic and clinical associations but not absolute agreement with reference Centiloid measurements. Next, the PLS-PVC point-spread function was fixed at 4.4 mm for all scanners rather than matching to each scanner’s resolution. Furthermore, the clinical analyses were cross-sectional and therefore characterize associations with clinical severity rather than progression. Finally, the study did not establish MUSE CL thresholds for amyloid positivity; because published thresholds depend on the criterion used to define positivity,^63–66^ such thresholds require appropriate amyloid reference standards. Future studies should thus evaluate the transformations across additional datasets, scanners, acquisition protocols and longitudinal cohorts, and establish MUSE CL thresholds for amyloid positivity.

## Data Availability

The imaging and clinical data analyzed in this study were obtained from the Alzheimer's Disease Neuroimaging Initiative (ADNI), the Charles F. and Joanne Knight Alzheimer Disease Research Center (Knight-ADRC), and the Centiloid reference and Navitsky datasets. The Knight-ADRC dataset is available at https://doi.org/10.17632/sf7gxzdn63. The Centiloid reference and Navitsky datasets were made available through the Global Alzheimer's Association Interactive Network (GAAIN; https://www.gaain.org). ADNI data are accessible through the ADNI data-sharing platform (https://adni.loni.usc.edu), subject to applicable data access requirements.

## Acknowledgements

Data collection and sharing for the Alzheimer’s Disease Neuroimaging Initiative (ADNI) is funded by the National Institute on Aging (National Institutes of Health Grant U19 AG024904). The grantee organization is the Northern California Institute for Research and Education. In the past, ADNI has also received funding from the National Institute of Biomedical Imaging and Bioengineering, the Canadian Institutes of Health Research, and private sector contributions through the Foundation for the National Institutes of Health (FNIH) including generous contributions from the following: AbbVie, Alzheimer’s Association; Alzheimer’s Drug Discovery Foundation; Araclon Biotech; BioClinica, Inc.; Biogen; Bristol-Myers Squibb Company; CereSpir, Inc.; Cogstate; Eisai Inc.; Elan Pharmaceuticals, Inc.; Eli Lilly and Company; EuroImmun; F. Hoffmann-La Roche Ltd and its affiliated company Genentech, Inc.; Fujirebio; GE Healthcare; IXICO Ltd.; Janssen Alzheimer Immunotherapy Research & Development, LLC.; Johnson & Johnson Pharmaceutical Research &Development LLC.; Lumosity; Lundbeck; Merck & Co., Inc.; Meso Scale Diagnostics, LLC.; NeuroRx Research; Neurotrack Technologies; Novartis Pharmaceuticals Corporation; Pfizer Inc.; Piramal Imaging; Servier; Takeda Pharmaceutical Company; and Transition Therapeutics.

The resources provided by the Charles F. and Joanne Knight Alzheimer Disease Research Center for doi: 10.17632/sf7gxzdn63 were supported by the National Institutes of Health under award numbers P30AG066444, P01AG03991, P01AG026276, R01AG078964, RF1AG044546, RF1AG071706, RF1AG058501, and P30NS048056. We extend our sincere gratitude to the Knight-ADRC research participants and their study partners for their invaluable time, contributions, and dedication to advancing this research. We also acknowledge the Knight-ADRC Cores and Projects for their essential efforts in conducting, curating, and disseminating these datasets.

We thank the original authors of the Centiloid Project for making the original Centiloid PiB reference dataset, including the young control and typical Alzheimer’s disease PET and MRI scans and the standard volumes of interest, publicly available through the Global Alzheimer’s Association Interactive Network (GAAIN; http://www.gaain.org). We also thank Michael Navitsky, Abhinay D. Joshi, Michael D. Devous Sr., and Avid Radiopharmaceuticals, a wholly owned subsidiary of Eli Lilly and Company, for making the paired [¹¹C]PiB and [¹⁸F]florbetapir dataset publicly available through GAAIN. These shared resources made the calibration reported here possible.

## Ethics declaration

Data used in this study were obtained from the Alzheimer’s Disease Neuroimaging Initiative (ADNI) and the Knight Alzheimer Disease Research Center (Knight ADRC) at Washington University in St. Louis. Institutional review board (IRB) approvals were obtained at each participating institution of these cohorts in accordance with the Declaration of Helsinki, and written informed consent was obtained from all participants or their authorized representatives before enrollment. The Washington University in St. Louis IRB approved the use of these existing data for the present study.

## Data availability

Datasets are available upon reasonable request following applicable guidelines of human subjects’ protection.

