## Supplementary Material for "Centiloid Calibration and Application of MUSE-Based Amyloid PET Quantification for [¹¹C]PiB and [¹⁸F]Florbetapir (AV-45)"

### Supplementary Methods

#### A. MRI and PET processing parameters

All MRI processing used MUSE 3.1.0; PET processing used a nipype-based pipeline with SPM12 (r7219), FSL 6.0.3, and a MATLAB implementation of PLS-PVC. Identical settings were used for all participants within each dataset. Sites applying the MUSE Centiloid transformations should reproduce these settings; departures require recalibration, as outlined in the main text.

| Step | Description | Software and key settings |
|---|---|---|
| Reorientation | T1 reoriented to LPS | N/A |
| Bias correction | Intensity non-uniformity correction | ANTs N4 |
| Skull stripping | Multi-atlas brain extraction; brain mask applied to the N4 image | MUSE 3.1.0, brain-extraction template set (50 templates); 15 best-matching templates registered; DRAMMS + ANTs registration (DRAMMS regularization 0.1, ANTs SyN 0.5); intensity correction and fuzzy CSF off |
| Second bias correction | Bias correction of the skull-stripped image | FSL FAST, 3 tissue classes, 8 bias-field iterations, 20 mm bias-field smoothing |
| Parcellation | Multi-atlas labeling into MUSE anatomical ROIs | MUSE 3.1.0, atlas set including the cerebellum (35 atlases); 11 most similar atlases registered; DRAMMS + ANTs registration (DRAMMS regularization 0.1, ANTs SyN 0.5); CSF weight 1.2; intensity-, similarity- and fuzzy-CSF-weighted label fusion (defaults); no white-matter lesion mask |

PET processing steps are outlined below.

| Step | Description | Software and key settings |
|---|---|---|
| Frame realignment (dynamic data) | Rigid motion correction of each frame to the mean of the early frames | SPM12 Realign: quality 1, 2 mm sampling, 7 mm smoothing, 2nd-degree B-spline (estimation), 4th-degree B-spline (reslicing) |
| Uptake-window averaging | Frame-duration-weighted mean image | Frames within the window (±0.5-min tolerance at the boundaries); Klunk PiB 50–70 min (static); Navitsky PiB 50–70 min (5-min frames); Navitsky AV-45 50–60 min (2 × 5 min); Knight-ADRC PiB 50–60 min (last 10 min of a 30–60 min acquisition); ADNI AV-45 50–70 min (4 × 5 min) |

| | | |
|---|---|---|
| Coregistration | Mean PET registered to the skull-stripped T1 | FSL FLIRT: 4 mm Gaussian pre-smoothing of the PET, 6 degrees of freedom, mutual information, ±30° search range |
| Labels to PET space | MUSE labels and T1 resampled into native PET space; labels restricted to the PET field of view | FSL FLIRT with the inverted transform; nearest-neighbour interpolation for labels |
| PLS-PVC (PLS-PVC runs only) | Parallel level set partial-volume correction with the T1 as anatomical prior | Isotropic PSF 4.4 mm FWHM; regularization weight 48; 100 iterations |
| SUVr | Voxelwise PET ÷ mean reference uptake; ROI means from MUSE labels and composites | Reference: MUSE labels 38/39 (cerebellar cortex). Target: mean cortical composite (**Supplementary Methods B**). Same label definitions for PiB and AV-45 |

#### B. MUSE global cortical ROI definition

The global cortical SUVr was calculated over the MUSE mean cortical composite, comprising the following 36 regions in each hemisphere: anterior cingulate gyrus, anterior orbital gyrus, angular gyrus, central operculum, frontal operculum, frontal pole, gyrus rectus, inferior occipital gyrus, inferior temporal gyrus, lateral orbital gyrus, middle cingulate gyrus, medial frontal cortex, middle frontal gyrus, middle occipital gyrus, medial orbital gyrus, superior frontal gyrus medial segment, middle temporal gyrus, occipital pole, opercular part of the inferior frontal gyrus, orbital part of the inferior frontal gyrus, posterior cingulate gyrus, precuneus, parietal operculum, posterior orbital gyrus, planum polare, planum temporale, subcallosal area, superior frontal gyrus, supplementary motor cortex, supramarginal gyrus, superior occipital gyrus, superior parietal lobule, superior temporal gyrus, temporal pole, triangular part of the inferior frontal gyrus, and transverse temporal gyrus.

#### C. Scanner effects in ADNI and harmonization sensitivity analysis

ComBat (neuroCombat v0.2.12) was applied with scanner family as the batch variable and diagnosis, age, and sex as protected covariates. The primary harmonized version was applied directly to global Centiloid values without empirical Bayes estimation, because empirical Bayes pooling requires multiple features. Two further variants were run. The first omitted diagnosis as a protected covariate, to check that improved separation was not imposed by the model. In the second, ComBat with empirical Bayes estimation was applied jointly to the global and predefined regional SUVrs, and the harmonized global SUVr was converted to Centiloid using the AV-45 equation. For each version, AD and CN means and SDs, Hedges' *g*, AUC, and the residual scanner share of variance were computed. Harmonization was performed on NoPVC Centiloid values only.

ComBat reduced the scanner-associated share of total Centiloid variance from 33.5% to below 0.1%, and Hedges' g increased from 0.892 to 1.116 and the AUC from 0.711 to 0.789 (Supplementary Table 11). Most of this gain came from reduced dispersion in the AD group, whose SD fell from 74.8 to 49.9 CL. Group means changed little: the AD mean went from 63.9 to 62.4 CL and the CN mean from 6.1 to 7.6 CL. The two alternative versions of ComBat gave similar results. Harmonization without diagnosis as a protected covariate yielded $g = 1.14$, and joint regional harmonization with empirical Bayes estimation yielded $g = 1.12$. Because diagnosis was included as a protected covariate, these improvements reflect in-sample harmonization effects rather than independently validated diagnostic performance.

However, ComBat did not resolve the substantial scanner-specific differences in absolute Centiloid values or diagnostic separation. Following ComBat adjustment for scanner-associated distributional differences, HR+ CN values increased from −34.5 to 36.7 CL and HR+ AD values increased from −20.2 to 33.1 CL. The poor AD–CN separation on the HR+ therefore remained after harmonization, with mean AD values slightly below CN values. CN values on the Siemens Biograph64/mCT (n = 2) moved from −89.1 to −44.8 CL and remained well below zero.

### Supplementary Figures

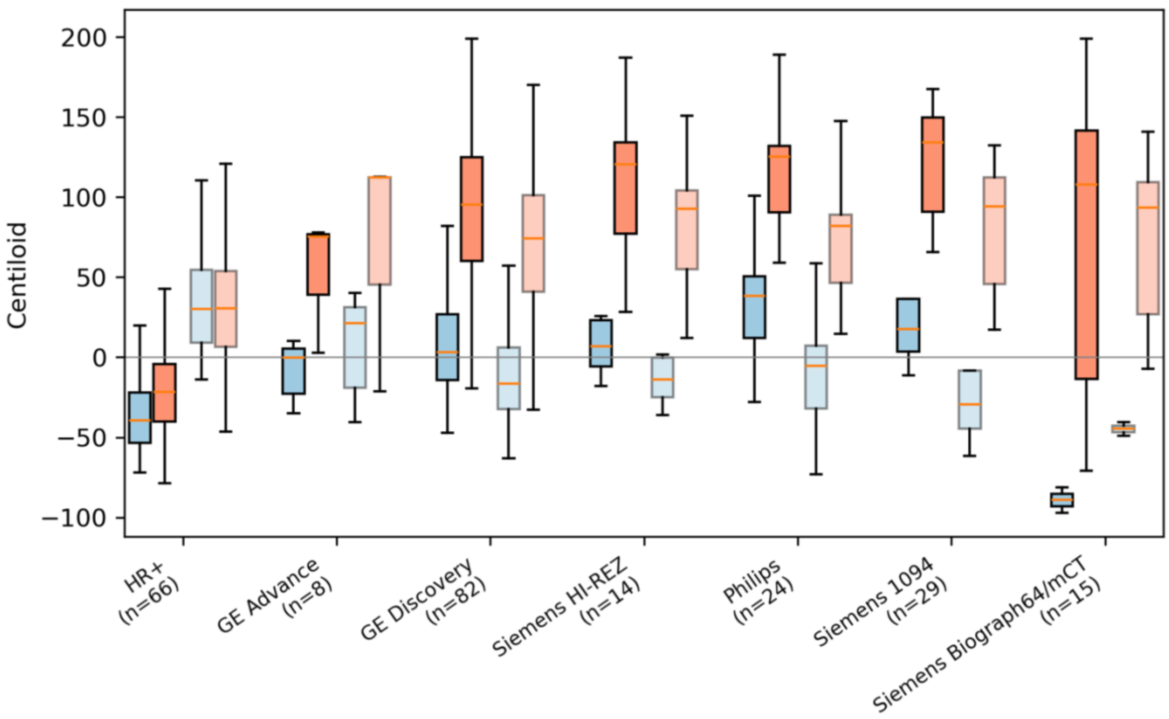

**Supplementary Figure 1. Centiloid values by scanner family and diagnosis in the ADNI AV-45 cohort (NoPVC), before and after ComBat harmonization.**

Box plots show Centiloid values for cognitively normal (CN, blue) and Alzheimer's disease (AD, red) participants within each scanner family. Solid boxes show unharmonized values. Faded boxes show values after ComBat harmonization with scanner family as the batch variable and diagnosis, age, and sex as protected covariates. Boxes span the interquartile range, the horizontal line marks the median, and whiskers extend to 1.5 × the interquartile range; outliers are not shown. Scanner families are ordered by median unharmonized Centiloid, and the number of participants in each family is given below its label. The horizontal grey line marks 0 CL.

### Supplementary Tables

#### Supplementary Table 1. PET and MRI acquisition and reconstruction parameters of the application cohorts, by PET scanner

| Cohort | PET scanner | N | Injected dose, MBq, median (range) | Frames | Reconstruction (N) | MRI scanner (N) | MRI field strength, T |
|---|---|---|---|---|---|---|---|
| Knight-ADRC PiB | Biograph mMR | 24 | 603 (292–725) | 6 × 5 min | OP-OSEM3i21s (23); OP-OSEM 3i 24s (1) | SIEMENS Biograph_mMR (23); SIEMENS Prisma_fit (1) | 3 |
| Knight-ADRC PiB | Biograph128 Vision 600 Edge | 6 | 544 (429–607) | 6 × 5 min | OSEM3D+TOF 8i5s (3); PSF+TOF 8i5s (2); Backprojection+TOF (1) | Siemens MAGNETOM Vida (5); SIEMENS Prisma_fit (1) | 3 |
| Knight-ADRC PiB | Biograph40 TruePoint | 3 | 585 (485–696) | 6 × 5 min | Backprojection | Siemens MAGNETOM Vida (2); SIEMENS Biograph_mMR (1) | 3 |
| ADNI AV-45 | GE Advance | 8 | 369 (344–397); not recorded or implausible in 4 | 4 × 5 min | not recorded | Philips Medical Systems Achieva (4); SIEMENS TrioTim (4) | 3 |
| ADNI AV-45 | GE Discovery | 82 | 358 (255–405); not recorded or implausible in 8 | 4 × 5 min | 3D IR (60); 3D FORE IR (18); VPHD (4) | SIEMENS TrioTim (22); SIEMENS Skyra (14); GE MEDICAL SYSTEMS DISCOVERY MR750 (13); SIEMENS Verio (12); Philips Medical Systems Achieva (10); GE MEDICAL SYSTEMS Signa HDxt (7); Philips Medical Systems GEMINI (2); Philips Medical Systems Ingenuity (1); Philips Healthcare Ingenia (1) | 3 |
| ADNI AV-45 | HR+ | 66 | 363 (259–407); not recorded or implausible in 19 | 4 × 5 min | osem-wa4/16 | SIEMENS TrioTim (25); GE MEDICAL SYSTEMS DISCOVERY MR750 (15); SIEMENS Skyra (8); Philips Medical Systems Ingenia (6); Philips Medical Systems Achieva (5); GE MEDICAL SYSTEMS Signa HDxt (4); GE MEDICAL SYSTEMS SIGNA HDx (3) | 3 |
| ADNI AV-45 | Philips | 24 | 351 (296–387) | 4 × 5 min | LOR-RAMLA | SIEMENS TrioTim (14); Philips Medical Systems Intera (3); GE MEDICAL SYSTEMS DISCOVERY MR750w (3); GE MEDICAL SYSTEMS Signa HDxt (2); GE MEDICAL SYSTEMS DISCOVERY MR750 (2) | 3 |
| ADNI AV-45 | Siemens 1094 | 29 | 354 (289–386) | 4 × 5 min | OSEM2D 4i16s | SIEMENS TrioTim (15); GE MEDICAL SYSTEMS Signa HDxt (14) | 3 |
| ADNI AV-45 | Siemens Biograph64/mCT | 15 | 349 (185–403) | 4 × 5 min | OSEM3D 4i12s (14); OSEM3D 4i24s (1) | Philips Medical Systems Achieva (14); SIEMENS Skyra (1) | 3 |
| ADNI AV-45 | Siemens HI-REZ | 14 | 370 (296–400) | 4 × 5 min | OSEM2D 4i16s (13); OSEM2D 4i14s (1) | GE MEDICAL SYSTEMS Signa HDxt | 3 |

Parameters were read from the DICOM (or ECAT 7, for the ECAT HR+) headers of the PET series processed in this study and of the corresponding T1-weighted MRI. N is the number of participants scanned on each PET scanner. Knight-ADRC PiB scans were acquired as six 5-min frames from 30 to 60 min after injection, and the last two frames (50–60 min) were averaged; ADNI AV-45 scans were acquired as four 5-min frames from 50 to 70 min after injection, and all frames were averaged. Reconstructed frames were used as provided by each site, and frame realignment was applied during processing. Injected dose is given as median (range); "not recorded or implausible" counts participants whose header contained no dose or a value outside 100–1000 MBq. Reconstruction abbreviations: OP-OSEM, ordinary Poisson ordered-subset expectation maximization; OSEM2D/OSEM3D, two- or three-dimensional OSEM; osem-wa, attenuation-weighted OSEM; *n*i*m*s, *n* iterations and *m* subsets; TOF, time-of-flight; PSF, point-spread-function modeling; 3D IR, three-dimensional iterative reconstruction; FORE, Fourier rebinning; VPHD, VUE Point HD; LOR-RAMLA, line-of-response row-action maximum-likelihood algorithm; Backprojection, filtered backprojection. Abbreviations: ADNI, Alzheimer's Disease Neuroimaging Initiative; ADRC, Alzheimer Disease Research Center; AV-45, [$^{18}$F]florbetapir; MRI, magnetic resonance imaging; PET, positron emission tomography; PiB, [$^{11}$C]Pittsburgh compound B.

**Supplementary Table 2. Anatomical composition of predefined MUSE regional gray matter targets**

| MUSE regional target | Constituent anatomical regions |
|---|---|
| **Posterior cingulate** | Bilateral posterior cingulate gyrus |
| **Lateral temporal** | Bilateral inferior temporal gyrus, middle temporal gyrus, superior temporal gyrus, and temporal pole |
| **Frontal** | Bilateral anterior and posterior insula; anterior, lateral, medial, and posterior orbital gyri; frontal pole; middle and superior frontal gyri; medial frontal cortex; gyrus rectus; precentral gyrus and its medial segment; superior frontal gyrus medial segment; supplementary motor cortex; subcallosal area; central, frontal, and parietal opercula; and opercular, orbital, and triangular parts of the inferior frontal gyrus |
| **Precuneus** | Bilateral precuneus |
| **Parietal** | Bilateral angular gyrus, postcentral gyrus and its medial segment, supramarginal gyrus, superior parietal lobule, and precuneus |
| **Occipital** | Bilateral calcarine cortex, cuneus, inferior, middle, and superior occipital gyri, occipital pole, occipital fusiform gyrus, and lingual gyrus |

Abbreviations: MUSE, MUlti-atlas region Segmentation utilizing Ensembles.

**Supplementary Table 3. Comparison of MUSE-derived and published Navitsky Centiloids (N = 43)**

| Tracer | Pearson *r* | $R^2$ | Slope | Intercept | Mean CL difference ± SD | ICC(A,1) (95% CI) |
|---|---|---|---|---|---|---|
| PiB | 0.986 ($P = 3.07 \times 10^{-33}$) | 0.971 | 1.20 | 1.5 | 8.6 ± 13.6 | 0.955 (0.868–0.980) |
| AV-45 | 0.987 ($P = 4.26 \times 10^{-34}$) | 0.974 | 1.25 | 0.1 | 9.0 ± 15.9 | 0.948 (0.866–0.976) |

Regression statistics characterize the relationship between published Navitsky and MUSE-derived CL measurements with NoPVC in the same 43 participants. Difference is MUSE – published; both mean differences were significant (paired t-test, $P < 0.001$). Abbreviations: AV-45, [$^{18}$F]florbetapir; CL, Centiloid; MUSE, MUlti-atlas region Segmentation utilizing Ensembles; PiB, [$^{11}$C]Pittsburgh compound-B; PLS-PVC, parallel level set regularization-based partial-volume correction; NoPVC, without partial-volume correction; ICC, intraclass correlation coefficient.

**Supplementary Table 4. Participant-level MUSE-derived PiB SUVr and Centiloid measurements in the Klunk calibration dataset (N = 79)**

| ID | Group | Published WC SUVr | Published WC CL | MUSE NoPVC SUVr | MUSE PLS-PVC SUVr | MUSE NoPVC CL | MUSE PLS-PVC CL |
|---|---|---|---|---|---|---|---|
| AD01 | AD | 2.1 | 102.3 | 2.012 | 2.128 | 106.723 | 109.990 |
| AD02 | AD | 2.102 | 102.5 | 1.934 | 1.996 | 97.767 | 96.267 |
| AD03 | AD | 2.416 | 131.9 | 2.481 | 2.644 | 160.664 | 163.846 |
| AD04 | AD | 2.288 | 119.9 | 2.150 | 2.235 | 122.603 | 121.130 |
| AD05 | AD | 2.138 | 105.8 | 2.009 | 2.081 | 106.430 | 105.104 |
| AD06 | AD | 2.143 | 106.3 | 1.947 | 2.029 | 99.286 | 99.659 |
| AD07 | AD | 2.195 | 111.2 | 2.195 | 2.296 | 127.763 | 127.577 |
| AD08 | AD | 2.022 | 94.9 | 1.958 | 2.046 | 100.528 | 101.490 |
| AD09 | AD | 2 | 92.8 | 1.768 | 1.810 | 78.726 | 76.831 |
| AD10 | AD | 2.266 | 117.8 | 2.000 | 2.060 | 105.317 | 102.948 |
| AD11 | AD | 2.121 | 104.2 | 1.931 | 2.001 | 97.374 | 96.784 |
| AD12 | AD | 2.05 | 97.5 | 1.981 | 2.060 | 103.162 | 102.933 |
| AD13 | AD | 2.112 | 103.4 | 2.109 | 2.213 | 117.936 | 118.929 |
| AD14 | AD | 2.023 | 95 | 1.993 | 2.072 | 104.536 | 104.159 |
| AD15 | AD | 1.653 | 60.3 | 1.690 | 1.779 | 69.707 | 73.603 |
| AD16 | AD | 1.626 | 57.8 | 1.578 | 1.619 | 56.817 | 56.898 |
| AD17 | AD | 1.813 | 75.3 | 1.721 | 1.771 | 73.315 | 72.726 |
| AD18 | AD | 2.072 | 99.6 | 1.600 | 1.628 | 59.345 | 57.900 |
| AD19 | AD | 2.179 | 109.6 | 1.999 | 2.085 | 105.309 | 105.546 |
| AD20 | AD | 2.155 | 107.4 | 1.974 | 2.062 | 102.352 | 103.134 |
| AD21 | AD | 2.39 | 129.5 | 2.188 | 2.294 | 126.940 | 127.327 |
| AD22 | AD | 2.347 | 125.4 | 2.252 | 2.354 | 134.365 | 133.613 |
| AD23 | AD | 2.105 | 102.7 | 2.004 | 2.085 | 105.875 | 105.556 |
| AD24 | AD | 2.175 | 109.3 | 1.919 | 1.987 | 96.080 | 95.300 |
| AD25 | AD | 1.668 | 61.7 | 1.639 | 1.674 | 63.838 | 62.656 |
| AD26 | AD | 2.113 | 103.4 | 1.996 | 2.085 | 104.899 | 105.510 |
| AD27 | AD | 2.315 | 122.4 | 2.007 | 2.069 | 106.152 | 103.891 |
| AD28 | AD | 2.2 | 111.6 | 2.278 | 2.434 | 137.305 | 141.945 |
| AD29 | AD | 1.984 | 91.4 | 1.816 | 1.892 | 84.154 | 85.356 |
| AD30 | AD | 2.27 | 118.2 | 2.050 | 2.136 | 111.095 | 110.835 |
| AD31 | AD | 1.826 | 76.6 | 1.784 | 1.833 | 80.469 | 79.233 |
| AD32 | AD | 1.938 | 87.1 | 1.971 | 2.077 | 102.005 | 104.677 |
| AD33 | AD | 1.896 | 83.1 | 1.727 | 1.788 | 73.982 | 74.505 |
| AD34 | AD | 2.04 | 96.6 | 2.074 | 2.178 | 113.823 | 115.233 |
| AD35 | AD | 1.874 | 81 | 1.764 | 1.819 | 78.183 | 77.796 |
| AD36 | AD | 2.183 | 110 | 2.106 | 2.161 | 117.586 | 113.494 |
| AD37 | AD | 2.007 | 93.5 | 1.701 | 1.739 | 70.959 | 69.486 |
| AD38 | AD | 2.233 | 114.7 | 2.287 | 2.411 | 138.428 | 139.560 |
| AD39 | AD | 1.888 | 82.4 | 1.798 | 1.872 | 82.180 | 83.311 |

| | | | | | | | |
|---|---|---|---|---|---|---|---|
| AD40 | AD | 2.183 | 110 | 2.064 | 2.147 | 112.753 | 112.026 |
| AD41 | AD | 1.635 | 58.7 | 1.538 | 1.571 | 52.270 | 51.954 |
| AD42 | AD | 2.405 | 130.9 | 2.080 | 2.137 | 114.615 | 110.922 |
| AD43 | AD | 2.259 | 117.1 | 2.192 | 2.324 | 127.408 | 130.454 |
| AD44 | AD | 1.907 | 84.1 | 1.825 | 1.889 | 85.288 | 85.098 |
| AD45 | AD | 2.11 | 103.2 | 1.812 | 1.867 | 83.687 | 82.807 |
| YC101 | YC | 0.964 | -4.3 | 1.045 | 1.022 | -4.429 | -5.359 |
| YC102 | YC | 1.008 | -0.1 | 1.045 | 1.024 | -4.458 | -5.120 |
| YC103 | YC | 0.947 | -5.8 | 1.052 | 1.059 | -3.679 | -1.503 |
| YC104 | YC | 1.048 | 3.6 | 1.071 | 1.066 | -1.484 | -0.785 |
| YC105 | YC | 1.057 | 4.4 | 1.030 | 1.024 | -6.162 | -5.131 |
| YC106 | YC | 1.08 | 6.6 | 1.133 | 1.128 | 5.657 | 5.701 |
| YC107 | YC | 1.116 | 10 | 1.202 | 1.186 | 13.531 | 11.737 |
| YC108 | YC | 1.048 | 3.6 | 1.149 | 1.132 | 7.486 | 6.100 |
| YC109 | YC | 0.997 | -1.1 | 1.064 | 1.042 | -2.249 | -3.304 |
| YC110 | YC | 1.023 | 1.3 | 1.103 | 1.085 | 2.225 | 1.241 |
| YC111 | YC | 1.026 | 1.6 | 1.104 | 1.092 | 2.306 | 1.988 |
| YC112 | YC | 1.093 | 7.9 | 1.145 | 1.138 | 7.048 | 6.721 |
| YC113 | YC | 0.977 | -3.1 | 1.040 | 1.026 | -5.015 | -4.937 |
| YC114 | YC | 1.026 | 1.6 | 1.114 | 1.109 | 3.466 | 3.693 |
| YC115 | YC | 0.964 | -4.3 | 1.061 | 1.049 | -2.588 | -2.489 |
| YC116 | YC | 0.974 | -3.4 | 1.035 | 1.031 | -5.632 | -4.379 |
| YC117 | YC | 0.963 | -4.3 | 1.105 | 1.109 | 2.406 | 3.733 |
| YC118 | YC | 0.975 | -3.2 | 0.985 | 0.976 | -11.415 | -10.088 |
| YC119 | YC | 1.024 | 1.4 | 1.133 | 1.129 | 5.641 | 5.808 |
| YC120 | YC | 1.013 | 0.3 | 1.073 | 1.053 | -1.232 | -2.139 |
| YC121 | YC | 0.984 | -2.4 | 1.066 | 1.052 | -2.081 | -2.185 |
| YC122 | YC | 0.973 | -3.4 | 1.090 | 1.073 | 0.668 | -0.005 |
| YC123 | YC | 0.951 | -5.5 | 1.034 | 1.027 | -5.784 | -4.859 |
| YC124 | YC | 0.974 | -3.3 | 1.093 | 1.085 | 1.084 | 1.178 |
| YC125 | YC | 1.047 | 3.5 | 1.075 | 1.072 | -1.028 | -0.163 |
| YC126 | YC | 0.959 | -4.7 | 1.050 | 1.033 | -3.882 | -4.162 |
| YC127 | YC | 1.032 | 2.1 | 1.085 | 1.076 | 0.160 | 0.272 |
| YC128 | YC | 0.937 | -6.8 | 1.004 | 1.001 | -9.161 | -7.558 |
| YC129 | YC | 1.002 | -0.7 | 1.092 | 1.085 | 0.975 | 1.272 |
| YC130 | YC | 0.976 | -3.1 | 1.102 | 1.100 | 2.041 | 2.833 |
| YC131 | YC | 1.039 | 2.7 | 1.127 | 1.107 | 4.943 | 3.547 |
| YC132 | YC | 1.093 | 7.9 | 1.122 | 1.111 | 4.348 | 3.961 |
| YC133 | YC | 0.988 | -2.1 | 1.105 | 1.083 | 2.475 | 1.021 |
| YC134 | YC | 1.045 | 3.3 | 1.117 | 1.105 | 3.819 | 3.359 |

Participant-level published reference and MUSE-derived PiB SUVr and Centiloid (CL) measurements are shown for the 79 participants in the Klunk calibration sample. Published CTX/WC SUVr and CL values are presented alongside MUSE-derived measurements obtained using PLS-PVC and NoPVC processing. MUSE-derived CL values were calculated using the corresponding MUSE-specific transformations described in the main manuscript. Negative CL values were retained without truncation. Abbreviations: AD, Alzheimer disease; CL, Centiloid; CTX/WC, cortical target-to-whole-cerebellum reference ratio; MUSE, MUlti-atlas region Segmentation utilizing Ensembles; PiB, [$^{11}$C]Pittsburgh compound B; PLS-PVC, parallel level set regularization-based partial-volume correction; SUVr, standardized uptake value ratio; NoPVC, without partial-volume correction; YC, young cognitively normal; WC, whole cerebellum.

**Supplementary Table 5. MUSE SUVr values and Centiloids for AV-45 calibration dataset.**

| ID | PiB NoPVC SUVr | AV-45 NoPVC SUVr | PiB PLS-PVC SUVr | AV-45 PLS-PVC SUVr | PiB NoPVC CL | AV-45 NoPVC CL | PiB PLS-PVC CL | AV-45 PLS-PVC CL |
|---|---|---|---|---|---|---|---|---|
| 105858 | 2.380 | 1.974 | 2.545 | 2.068 | 149.1 | 182.5 | 153.5 | 189.1 |
| 106843 | 1.112 | 1.168 | 1.101 | 1.152 | 3.2 | 6.5 | 2.9 | 6.9 |
| 123723 | 0.947 | 1.081 | 0.948 | 1.081 | -15.7 | -12.3 | -13.0 | -7.3 |
| 123950 | 1.721 | 1.452 | 1.768 | 1.445 | 73.3 | 68.6 | 72.4 | 65.2 |
| 130779 | 1.110 | 1.152 | 1.109 | 1.152 | 3.0 | 3.0 | 3.8 | 6.9 |
| 146743 | 2.318 | 1.834 | 2.454 | 1.878 | 142.0 | 151.8 | 144.1 | 151.2 |
| 164067 | 2.112 | 1.469 | 2.178 | 1.481 | 118.3 | 72.3 | 115.2 | 72.3 |
| 168001 | 2.100 | 1.767 | 2.171 | 1.789 | 116.9 | 137.2 | 114.5 | 133.6 |
| 181120 | 1.083 | 1.159 | 1.080 | 1.142 | -0.1 | 4.6 | 0.7 | 4.9 |
| 196851 | 1.176 | 1.171 | 1.171 | 1.152 | 10.5 | 7.2 | 10.2 | 6.9 |
| 214174 | 1.145 | 1.154 | 1.141 | 1.145 | 7.1 | 3.5 | 7.1 | 5.4 |
| 225403 | 2.099 | 1.637 | 2.187 | 1.649 | 116.7 | 109.0 | 116.2 | 105.6 |
| 242877 | 1.918 | 1.709 | 2.013 | 1.754 | 95.9 | 124.7 | 98.0 | 126.7 |
| 271648 | 1.100 | 1.229 | 1.094 | 1.209 | 1.8 | 19.9 | 2.2 | 18.1 |
| 291467 | 1.876 | 1.390 | 1.937 | 1.394 | 91.1 | 54.9 | 90.1 | 55.0 |
| 293755 | 1.263 | 1.409 | 1.262 | 1.403 | 20.6 | 59.2 | 19.7 | 56.7 |
| 303610 | 1.123 | 1.281 | 1.105 | 1.248 | 4.5 | 31.3 | 3.3 | 25.9 |
| 313017 | 1.962 | 1.759 | 2.023 | 1.803 | 101.0 | 135.5 | 99.0 | 136.3 |
| 335214 | 1.100 | 1.016 | 1.093 | 1.002 | 1.9 | -26.6 | 2.1 | -23.0 |
| 346724 | 0.947 | 1.027 | 0.948 | 1.015 | -15.8 | -24.1 | -13.1 | -20.6 |
| 370972 | 2.073 | 1.657 | 2.156 | 1.669 | 113.8 | 113.3 | 113.0 | 109.8 |
| 378784 | 2.250 | 1.444 | 2.384 | 1.488 | 134.1 | 66.9 | 136.8 | 73.7 |
| 388397 | 1.087 | 1.107 | 1.073 | 1.090 | 0.3 | -6.6 | -0.1 | -5.5 |
| 395069 | 1.914 | 1.623 | 1.989 | 1.646 | 95.5 | 105.8 | 95.5 | 105.1 |
| 422325 | 2.521 | 1.807 | 2.696 | 1.870 | 165.2 | 146.0 | 169.3 | 149.6 |
| 444992 | 2.489 | 2.068 | 2.603 | 2.121 | 161.6 | 202.9 | 159.6 | 199.8 |
| 457754 | 1.527 | 1.312 | 1.553 | 1.306 | 51.0 | 37.9 | 50.0 | 37.4 |
| 461664 | 1.217 | 1.053 | 1.220 | 1.041 | 15.3 | -18.6 | 15.3 | -15.3 |
| 466541 | 1.919 | 1.581 | 2.022 | 1.623 | 96.0 | 96.6 | 98.9 | 100.5 |
| 476741 | 1.432 | 1.273 | 1.453 | 1.267 | 40.0 | 29.4 | 39.6 | 29.8 |
| 101437 | 1.119 | 1.162 | 1.115 | 1.145 | 4.0 | 5.3 | 4.4 | 5.5 |
| 119551 | 1.108 | 1.204 | 1.102 | 1.178 | 2.7 | 14.4 | 3.0 | 12.0 |
| 198665 | 1.016 | 1.071 | 1.004 | 1.053 | -7.8 | -14.6 | -7.2 | -12.9 |
| 296898 | 1.042 | 1.086 | 1.044 | 1.062 | -4.8 | -11.4 | -3.1 | -11.2 |
| 315554 | 1.079 | 1.112 | 1.063 | 1.088 | -0.6 | -5.6 | -1.0 | -5.9 |
| 363785 | 1.095 | 1.143 | 1.091 | 1.127 | 1.3 | 1.2 | 1.9 | 1.9 |
| 395690 | 1.036 | 1.121 | 1.034 | 1.100 | -5.5 | -3.7 | -4.1 | -3.6 |
| 404333 | 1.165 | 1.166 | 1.157 | 1.138 | 9.3 | 6.1 | 8.8 | 4.0 |
| 406808 | 1.099 | 1.125 | 1.088 | 1.097 | 1.8 | -2.7 | 1.5 | -4.2 |

| | | | | | | | | |
|---|---|---|---|---|---|---|---|---|
| 411884 | 1.079 | 1.118 | 1.076 | 1.111 | -0.5 | -4.3 | 0.3 | -1.3 |
| 453954 | 1.144 | 1.325 | 1.143 | 1.306 | 6.9 | 40.9 | 7.3 | 37.4 |
| 462623 | 1.053 | 1.140 | 1.038 | 1.114 | -3.6 | 0.5 | -3.7 | -0.8 |
| 487972 | 1.180 | 1.155 | 1.167 | 1.133 | 11.1 | 3.9 | 9.8 | 3.1 |

MUSE-based SUVr and CL values are presented using PLS-PVC and NoPVC processing. MUSE-derived CL values were calculated using the corresponding MUSE-specific transformations described in the main manuscript. Negative CL values were retained without truncation. Abbreviations: AD, Alzheimer disease; CL, Centiloid; MUSE, MUlti-atlas region Segmentation utilizing Ensembles; PLS-PVC, parallel level set regularization-based partial-volume correction; SUVr, standardized uptake value ratio; NoPVC, without partial-volume correction; YC, young cognitively normal.

#### Supplementary Table 6. Characteristics of the independent AV-45 and PiB application cohorts

| Cohort | Characteristic | AD | CN | P |
|---|---|---|---|---|
| ADNI AV-45 | N | 119 | 119 | — |
| | Age | 73.66 ± 8.08 | 73.80 ± 6.25 | 0.882 |
| | Education | 15.82 ± 2.76 | 16.28 ± 2.44 | 0.172 |
| | MMSE | 23.11 ± 2.10 | 29.07 ± 1.21 | $3.60 \times 10^{-66}$ |
| | CDR-SB | 4.54 ± 1.64 | 0.03 ± 0.11 | $2.74 \times 10^{-45}$ |
| | CDR | 0.81 ± 0.27 | 0.00 ± 0.05 | $1.92 \times 10^{-47}$ |
| | Female, % | 43.7 | 48.7 | 0.516 |
| Knight-ADRC PiB | N | 18 | 15 | — |
| | Age | 72.44 ± 5.43 | 66.00 ± 6.98 | 0.007 |
| | Education | 15.39 ± 2.68 | 16.27 ± 2.49 | 0.338 |
| | MMSE | 24.72 ± 4.64 | 29.50 ± 0.65 | $4.22 \times 10^{-4}$ |
| | CDR-SB | 2.94 ± 1.96 | 0.00 ± 0.00 | $3.05 \times 10^{-7}$ |
| | CDR | 0.61 ± 0.21 | 0.00 ± 0.00 | $9.20 \times 10^{-8}$ |
| | Female, % | 44.4 | 46.7 | 1.000 |

Values are presented as mean ± SD where available or counts, or percentages, as indicated. In ADNI AV-45, participants with AD dementia and CN participants were matched 1:1 and balanced for age, education, and sex. For the Knight-ADRC PiB application, participants with clinical assessments within 180 days of PET were included. MMSE and CDR-SB were compared with Welch's t-test and the Mann–Whitney U test, respectively. Sex was compared with the chi-squared test. Years of education and age were compared with Welch's t-test. In Knight-ADRC PiB, MMSE was available for 18 AD and 14 CN participants. Abbreviations: AD, Alzheimer disease; ADRC, Alzheimer Disease Research Center; ADNI, Alzheimer's Disease Neuroimaging Initiative; AV-45, [$^{18}$F]florbetapir; CN, cognitively normal; PET, positron emission tomography; PiB, [$^{11}$C]Pittsburgh compound-B; SD, standard deviation.

**Supplementary Table 7. Independent application of MUSE-derived PiB Centiloid measurements in the Knight-ADRC cohort**

| | NoPVC | PLS-PVC | PLS-PVC − NoPVC difference (95% CI) |
|---|---|---|---|
| AD dementia, CL, mean (SD) | 95.2 (66.5) | 101.2 (71.4) | — |
| CN, CL, mean (SD) | –2.6 (13.2) | –2.0 (12.6) | — |
| AD−CN mean difference, CL | 97.8 | 103.2 | — |
| Hedges' *g* | 1.907 | 1.880 | –0.027 (–0.122, 0.023) |
| Welch's *P* | $8.04 \times 10^{-6}$ | $1.02 \times 10^{-5}$ | |
| CDR-SB, adjusted *ρ* | 0.536 | 0.512 | –0.024 (–0.084, 0.003) |
| Global CDR, adjusted *ρ* | 0.533 | 0.508 | –0.025 (–0.083, 0.004) |
| MMSE, adjusted *ρ* | –0.392 | –0.361 | 0.031 (–0.000, 0.114) |

Values are presented as mean ± SD unless otherwise indicated. Diagnostic separation was assessed between participants with AD dementia (N = 18) and CN participants (N = 15). Clinical associations are partial Spearman's correlations adjusted for age, sex, and education. Between-method differences in Hedges' g and adjusted Spearman's correlations (ρ) were evaluated using paired subject-level bootstrap resampling with 10,000 repetitions and percentile 95% confidence intervals. Between-method differences are defined as PLS-PVC − NoPVC. Abbreviations: AD, Alzheimer disease; CDR, Clinical Dementia Rating; CDR-SB, Clinical Dementia Rating Sum of Boxes; CN, cognitively normal; CL, Centiloid; MMSE, Mini-Mental State Examination; MUSE, MUlti-atlas region Segmentation utilizing Ensembles; PiB, [$^{11}$C]Pittsburgh compound B; PLS-PVC, parallel level set regularization-based partial-volume correction; NoPVC, without partial-volume correction.

**Supplementary Table 8. Independent application of MUSE-derived AV-45 Centiloid measurements in ADNI**

| | **NoPVC** | **PLS-PVC** | **PLS-PVC − NoPVC difference (95% CI)** |
|---|---|---|---|
| AD dementia, CL, mean (SD) | 63.9 (74.8) | 68.8 (74.0) | — |
| CN, CL, mean (SD) | 6.1 (52.6) | 8.9 (50.4) | — |
| AD−CN mean difference, CL | 57.864 | 59.987 | — |
| Hedges' *g* | 0.892 | 0.944 | 0.053 (0.038, 0.068) |
| Welch's *P* | $5.93 \times 10^{-11}$ | $5.71 \times 10^{-12}$ | — |
| CDR-SB, adjusted *ρ* | 0.334 | 0.356 | 0.022 (0.013, 0.032) |
| Global CDR, adjusted *ρ* | 0.372 | 0.394 | 0.022 (0.013, 0.032) |
| MMSE, adjusted *ρ* | –0.364 | –0.387 | –0.022 (–0.034, –0.013) |

Values are presented as mean ± SD unless otherwise indicated. Diagnostic separation was assessed between participants with AD dementia (N = 119) and CN participants (N = 119). Clinical associations are partial Spearman's correlations adjusted for age, sex, and education. Between-method differences in Hedges' *g* and adjusted Spearman's correlations *ρ* were evaluated using paired subject-level bootstrap resampling with 10,000 repetitions and percentile 95% confidence intervals. Between-method differences are defined as PLS-PVC − NoPVC. Abbreviations: AD, Alzheimer disease; ADNI, Alzheimer's Disease Neuroimaging Initiative; AV-45, [$^{18}$F]florbetapir; CDR, Clinical Dementia Rating; CDR-SB, Clinical Dementia Rating Sum of Boxes; CL, Centiloid; CN, cognitively normal; MMSE, Mini-Mental State Examination; MUSE, MUlti-atlas region Segmentation utilizing Ensembles; PLS-PVC, parallel level set regularization-based partial-volume correction; NoPVC, without partial-volume correction.

#### Supplementary Table 9. ADNI AV-45 application cohort by scanner

| Method | Scanner | AD N | CN N | AD mean CL (SD) | CN mean CL (SD) | AD-CN difference CL | Hedges *g* |
|---|---|---|---|---|---|---|---|
| NoPVC | GE Advance | 3 | 5 | 52.0 (42.7) | –8.5 (19.5) | 60.5 | 1.791 |
| | GE Discovery | 39 | 43 | 89.6 (53.2) | 20.8 (54.9) | 68.7 | 1.259 |
| | HR+ | 33 | 33 | –20.2 (28.3) | –34.5 (25.4) | 14.3 | 0.525 |
| | Philips | 9 | 15 | 124.8 (52.0) | 38.7 (43.4) | 86.0 | 1.777 |
| | Siemens 1094 | 14 | 15 | 123.6 (32.9) | 34.3 (49.4) | 89.3 | 2.055 |
| | Siemens Biograph64/mCT | 13 | 2 | 74.5 (90.0) | –89.1 (11.3) | 163.6 | 1.780 |
| | Siemens HI-REZ | 8 | 6 | 100.6 (72.0) | 15.1 (33.9) | 85.4 | 1.351 |
| | All except HR+ | 86 | 86 | 96.2 (60.6) | 21.6 (52.1) | 74.6 | 1.314 |
| PLS-PVC | GE Advance | 3 | 5 | 53.6 (43.9) | –6.9 (17.7) | 60.6 | 1.807 |
| | GE Discovery | 39 | 43 | 91.2 (52.0) | 21.8 (53.7) | 69.4 | 1.300 |
| | HR+ | 33 | 33 | –13.2 (26.3) | –27.1 (23.4) | 13.8 | 0.550 |
| | Philips | 9 | 15 | 130.5 (54.3) | 38.7 (43.2) | 91.8 | 1.865 |
| | Siemens 1094 | 14 | 15 | 132.5 (35.2) | 35.1 (52.2) | 97.4 | 2.112 |
| | Siemens Biograph64/mCT | 13 | 2 | 80.5 (89.0) | –78.1 (9.4) | 158.6 | 1.746 |
| | Siemens HI-REZ | 8 | 6 | 104.2 (73.3) | 15.4 (34.3) | 88.8 | 1.382 |
| | All except HR+ | 86 | 86 | 100.3 (61.0) | 22.6 (51.3) | 77.7 | 1.372 |

Rows above “All except HR+” are the seven scanner families; “All except HR+” pools the six non-HR+ families. Hedges’ *g* compares AD with CN within each row. The Siemens Biograph64/mCT row includes only two CN participants. AD, Alzheimer disease; CL, Centiloid; CN, cognitively normal; HR+, Siemens ECAT HR+; NoPVC, without partial-volume correction; PLS-PVC, parallel level set regularization-based partial-volume correction.

#### Supplementary Table 10. Knight-ADRC PiB application cohort by scanner

| Method | Scanner | Group | N | CL mean (SD) |
|---|---|---|---|---|
| NoPVC | Biograph128_Vision 600 Edge | AD | 6 | 92.9 (88.1) |
| | Biograph40_TruePoint | AD | 1 | 100.1 |
| | Biograph40_TruePoint | CN | 2 | –4.9 (7.8) |
| | Biograph_mMR | AD | 11 | 96.1 (60.3) |
| | Biograph_mMR | CN | 13 | –2.2 (14.0) |
| PLS-PVC | Biograph128_Vision 600 Edge | AD | 6 | 101.4 (96.9) |
| | Biograph40_TruePoint | AD | 1 | 103.1 |
| | Biograph40_TruePoint | CN | 2 | –4.2 (7.6) |
| | Biograph_mMR | AD | 11 | 100.9 (63.1) |
| | Biograph_mMR | CN | 13 | –1.7 (13.4) |

CL values are NoPVC and PLS-PVC global Centiloids. Within-scanner AD–CN comparisons are not reported because only the Biograph mMR had at least two participants in both diagnostic groups. AD, Alzheimer disease; CL, Centiloid; CN, cognitively normal; NoPVC, without partial-volume correction; PLS-PVC, parallel level set regularization-based partial-volume correction.

**Supplementary Table 11. Harmonization sensitivity analysis of NoPVC Centiloid in the ADNI AV-45 cohort**

| | Unharmonized | ComBat | ComBat without diagnosis | Regional ComBat (empirical Bayes) |
|---|---|---|---|---|
| AD dementia, CL, mean (SD) | 63.9 (74.8) | 62.4 (49.9) | 63.4 (51.5) | 62.7 (50.2) |
| CN, CL, mean (SD) | 6.1 (52.6) | 7.6 (48.1) | 6.6 (47.7) | 7.3 (48.2) |
| Hedges' *g* | 0.892 | 1.116 | 1.141 | 1.124 |
| AUC | 0.711 | 0.789 | 0.795 | 0.790 |
| Scanner share of CL variance, % | 33.5 | <0.1 | 1.2 | <0.1 |

Values are presented as mean (SD) unless otherwise indicated (119 AD dementia and 119 CN participants). ComBat used scanner family (seven families) as the batch variable and protected age, sex, and diagnosis (except where stated). ComBat and ComBat without diagnosis were applied to global Centiloid without empirical Bayes estimation; regional ComBat was applied with empirical Bayes estimation to the global and six predefined regional SUVr values, and the harmonized global SUVr was converted to Centiloid. Scanner share is the type II sum of squares for scanner family, from a model also including diagnosis, age, and sex, divided by the total sum of squares. Abbreviations: AD, Alzheimer disease; ADNI, Alzheimer's Disease Neuroimaging Initiative; AUC, area under the receiver operating characteristic curve; AV-45, [$^{18}$F]florbetapir; CL, Centiloid; CN, cognitively normal; NoPVC, without partial-volume correction; SUVr, standardized uptake value ratio.

#### Supplementary Table 12. Regional PiB–AV-45 cross-tracer regression parameters

| Gray matter target region | Cross-tracer slope PLS-PVC | Cross-tracer intercept PLS-PVC | Cross-tracer slope NoPVC | Cross-tracer intercept NoPVC | PiB CL equation PLS-PVC | AV-45 CL equation PLS-PVC | Cross-tracer $R^2$ PLS-PVC |
|---|---|---|---|---|---|---|---|
| Posterior cingulate | 0.501 | 0.573 | 0.505 | 0.617 | 102.1 × SUVr -122.3 | 203.8 × SUVr -239.1 | 0.814 |
| Lateral temporal GM | 0.526 | 0.533 | 0.530 | 0.540 | 107.7 × SUVr -110.6 | 204.8 × SUVr -219.8 | 0.883 |
| Frontal GM | 0.542 | 0.546 | 0.542 | 0.560 | 110.0 × SUVr -119.1 | 203.0 × SUVr -230.0 | 0.871 |
| Precuneus | 0.505 | 0.554 | 0.511 | 0.575 | 83.4 × SUVr -94.4 | 165.0 × SUVr -185.8 | 0.865 |
| Parietal GM | 0.523 | 0.567 | 0.525 | 0.576 | 107.2 × SUVr -117.1 | 204.7 × SUVr -233.2 | 0.864 |
| Occipital GM | 0.554 | 0.507 | 0.564 | 0.513 | 175.4 × SUVr -196.6 | 316.5 × SUVr -356.9 | 0.831 |

Cross-tracer parameters were obtained from region-specific linear regressions relating AV-45 SUVr to PiB SUVr. GM, gray matter; PiB, [$^{11}$C]Pittsburgh compound B; AV-45, [$^{18}$F]florbetapir; PLS-PVC, parallel level set regularization-based partial-volume correction; SUVr, standardized uptake value ratio; NoPVC, without partial-volume correction.

**Supplementary Table 13. MUSE-derived Centiloid calibration and PiB–AV-45 cross-tracer regression parameters for individual gray matter ROIs**

| Gray matter target region | PiB CL equation PLS-PVC | PiB CL equation NoPVC | AV-45 CL equation PLS-PVC | AV-45 CL equation NoPVC | Cross-tracer slope PLS-PVC | Cross-tracer intercept PLS-PVC | Cross-tracer $R^2$ PLS-PVC | Cross-tracer slope NoPVC | Cross-tracer intercept NoPVC | Cross-tracer $R^2$ NoPVC |
|---|---|---|---|---|---|---|---|---|---|---|
| Mean Cortical | 104.3 × SUVr -111.9 | 115.0 × SUVr -124.7 | 199.0 × SUVr -222.5 | 218.1 × SUVr -248.1 | 0.524 | 0.555 | 0.884 | 0.527 | 0.566 | 0.873 |
| Right Accumbens Area | 76.6 × SUVr -81.7 | 84.1 × SUVr -94.0 | 183.2 × SUVr -206.7 | 194.8 × SUVr -229.7 | 0.418 | 0.682 | 0.731 | 0.431 | 0.697 | 0.734 |
| Left Accumbens Area | 75.1 × SUVr -78.5 | 82.2 × SUVr -89.8 | 184.1 × SUVr -206.5 | 191.9 × SUVr -225.4 | 0.408 | 0.695 | 0.716 | 0.428 | 0.706 | 0.712 |
| Right Amygdala | 258.9 × SUVr -263.6 | 279.4 × SUVr -288.6 | 432.4 × SUVr -440.5 | 455.5 × SUVr -472.4 | 0.599 | 0.409 | 0.532 | 0.613 | 0.403 | 0.534 |
| Left Amygdala | 284.2 × SUVr -293.9 | 304.7 × SUVr -320.3 | 572.7 × SUVr -595.0 | 590.3 × SUVr -622.3 | 0.496 | 0.526 | 0.484 | 0.516 | 0.512 | 0.477 |
| Right Caudate | 134.3 × SUVr -145.2 | 155.3 × SUVr -172.9 | 342.2 × SUVr -396.1 | 390.3 × SUVr -464.0 | 0.392 | 0.733 | 0.573 | 0.398 | 0.746 | 0.534 |
| Left Caudate | 152.2 × SUVr -166.3 | 173.6 × SUVr -193.5 | 400.0 × SUVr -452.8 | 448.3 × SUVr -518.7 | 0.381 | 0.716 | 0.535 | 0.387 | 0.725 | 0.514 |
| Right Hippocampus | 970.2 × SUVr -1137.7 | 1049.3 × SUVr -1262.0 | 1698.1 × SUVr -1967.5 | 1915.2 × SUVr -2286.5 | 0.571 | 0.489 | 0.282 | 0.548 | 0.535 | 0.269 |
| Left Hippocampus | 1202.3 × SUVr -1447.3 | 1372.3 × SUVr -1695.2 | 2093.0 × SUVr -2441.8 | 2487.3 × SUVr -2981.4 | 0.574 | 0.475 | 0.292 | 0.552 | 0.517 | 0.279 |
| Right Pallidum | 352.3 × SUVr -562.2 | 305.8 × SUVr -478.3 | 1452.7 × SUVr -2501.9 | 1197.9 × SUVr -2029.4 | 0.243 | 1.335 | 0.152 | 0.255 | 1.295 | 0.175 |
| Left Pallidum | 301.5 × SUVr -474.9 | 271.1 × SUVr -417.0 | 1504.8 × SUVr -2610.9 | 1242.7 × SUVr -2105.3 | 0.200 | 1.419 | 0.113 | 0.218 | 1.359 | 0.147 |
| Right Putamen | 103.1 × SUVr -126.3 | 112.5 × SUVr -143.4 | 212.1 × SUVr -282.4 | 231.7 × SUVr -322.6 | 0.486 | 0.736 | 0.765 | 0.485 | 0.773 | 0.741 |
| Left Putamen | 104.0 × SUVr -127.1 | 113.2 × SUVr -143.8 | 243.0 × SUVr -324.0 | 265.7 × SUVr -368.0 | 0.428 | 0.810 | 0.707 | 0.426 | 0.844 | 0.684 |
| Right Thalamus Proper | 303.9 × SUVr -427.9 | 347.7 × SUVr -487.9 | 728.2 × SUVr -1038.7 | 801.2 × SUVr -1133.0 | 0.417 | 0.839 | 0.397 | 0.434 | 0.805 | 0.389 |
| Left Thalamus Proper | 360.7 × SUVr -525.5 | 415.7 × SUVr -601.7 | 886.9 × SUVr -1303.3 | 975.6 × SUVr -1415.2 | 0.407 | 0.877 | 0.336 | 0.426 | 0.834 | 0.334 |
| Cerebellar Vermal Lobules I-V | -1563.6 × SUVr +1557.9 | -1667 × SUVr +1661.5 | -4472.1 × SUVr +4463.1 | -4954.1 × SUVr +4885.2 | 0.350 | 0.650 | 0.169 | 0.336 | 0.651 | 0.144 |
| Cerebellar Vermal Lobules VI-VII | -1010.2 × SUVr +1039 | -1040.7 × SUVr +1071.7 | -3121.7 × SUVr +3090.7 | -3173.5 × SUVr +3123.4 | 0.324 | 0.657 | 0.202 | 0.328 | 0.647 | 0.166 |
| Cerebellar Vermal Lobules VIII-X | 17760.1 × SUVr -18345.5 | -40318 × SUVr +41968.1 | 42072.2 × SUVr -44574.7 | -84067.8 × SUVr +89179.5 | 0.422 | 0.623 | 0.294 | 0.480 | 0.562 | 0.337 |
| Left Basal Forebrain | 177.0 × SUVr -181.5 | 177.8 × SUVr -183.1 | 379.1 × SUVr -339.7 | 348.7 × SUVr -317.7 | 0.467 | 0.417 | 0.636 | 0.510 | 0.386 | 0.623 |
| Right Basal Forebrain | 175.7 × SUVr -187.4 | 174.1 × SUVr -185.5 | 369.5 × SUVr -352.0 | 342.3 × SUVr -324.7 | 0.476 | 0.445 | 0.619 | 0.509 | 0.407 | 0.619 |
| Right anterior cingulate gyrus | 77.8 × SUVr -87.4 | 83.6 × SUVr -97.0 | 165.3 × SUVr -190.2 | 176.2 × SUVr -212.1 | 0.471 | 0.622 | 0.849 | 0.474 | 0.653 | 0.838 |
| Left anterior cingulate gyrus | 76.6 × SUVr -85.7 | 82.1 × SUVr -94.7 | 161.9 × SUVr -186.2 | 175.0 × SUVr -210.2 | 0.473 | 0.621 | 0.847 | 0.469 | 0.660 | 0.836 |

| | | | | | | | | | | |
|---|---|---|---|---|---|---|---|---|---|---|
| Right anterior insula | 108.2 × SUVr -117.7 | 115.8 × SUVr -128.8 | 197.2 × SUVr -213.0 | 210.6 × SUVr -233.4 | 0.549 | 0.483 | 0.830 | 0.550 | 0.497 | 0.825 |
| Left anterior insula | 106.7 × SUVr -112.2 | 113.3 × SUVr -122.1 | 211.3 × SUVr -219.6 | 221.5 × SUVr -236.4 | 0.505 | 0.508 | 0.835 | 0.512 | 0.516 | 0.834 |
| Right anterior orbital gyrus | 110.8 × SUVr -121.3 | 122.5 × SUVr -136.2 | 205.3 × SUVr -244.1 | 223.1 × SUVr -274.2 | 0.540 | 0.599 | 0.832 | 0.549 | 0.619 | 0.820 |
| Left anterior orbital gyrus | 109.5 × SUVr -118.1 | 121.1 × SUVr -132.6 | 209.7 × SUVr -248.9 | 229.9 × SUVr -282.0 | 0.522 | 0.624 | 0.853 | 0.527 | 0.650 | 0.851 |
| Right angular gyrus | 88.0 × SUVr -94.2 | 97.1 × SUVr -104.8 | 169.3 × SUVr -193.4 | 186.2 × SUVr -216.2 | 0.520 | 0.586 | 0.861 | 0.521 | 0.598 | 0.857 |
| Left angular gyrus | 92.7 × SUVr -99.6 | 102.0 × SUVr -110.6 | 192.2 × SUVr -220.2 | 210.6 × SUVr -244.5 | 0.482 | 0.627 | 0.866 | 0.484 | 0.636 | 0.858 |
| Right calcarine cortex | 167.6 × SUVr -190.8 | 180.8 × SUVr -212.1 | 267.2 × SUVr -314.5 | 289.0 × SUVr -357.7 | 0.627 | 0.463 | 0.798 | 0.626 | 0.504 | 0.787 |
| Left calcarine cortex | 174.5 × SUVr -200.8 | 187.0 × SUVr -220.3 | 278.9 × SUVr -335.7 | 302.8 × SUVr -380.2 | 0.626 | 0.483 | 0.858 | 0.618 | 0.528 | 0.848 |
| Right central operculum | 118.1 × SUVr -133.4 | 130.2 × SUVr -152.1 | 194.5 × SUVr -227.6 | 213.9 × SUVr -264.2 | 0.607 | 0.484 | 0.815 | 0.609 | 0.524 | 0.792 |
| Left central operculum | 117.6 × SUVr -130.1 | 128.4 × SUVr -147.6 | 222.1 × SUVr -249.9 | 238.7 × SUVr -284.7 | 0.529 | 0.539 | 0.824 | 0.538 | 0.575 | 0.802 |
| Right cuneus | 231.3 × SUVr -271.5 | 242.1 × SUVr -288.6 | 420.8 × SUVr -481.9 | 432.5 × SUVr -509.7 | 0.550 | 0.500 | 0.721 | 0.560 | 0.511 | 0.709 |
| Left cuneus | 242.4 × SUVr -287.0 | 253.5 × SUVr -303.2 | 468.5 × SUVr -542.8 | 498.4 × SUVr -591.1 | 0.517 | 0.546 | 0.732 | 0.509 | 0.578 | 0.708 |
| Right entorhinal area | 391.2 × SUVr -426.8 | 429.3 × SUVr -472.9 | 632.5 × SUVr -600.6 | 697.8 × SUVr -666.0 | 0.618 | 0.275 | 0.490 | 0.615 | 0.277 | 0.478 |
| Left entorhinal area | 414.2 × SUVr -437.6 | 440.0 × SUVr -467.1 | 703.4 × SUVr -655.2 | 747.6 × SUVr -693.5 | 0.589 | 0.309 | 0.646 | 0.589 | 0.303 | 0.599 |
| Right frontal operculum | 94.8 × SUVr -104.6 | 103.2 × SUVr -118.5 | 172.0 × SUVr -199.0 | 183.9 × SUVr -224.0 | 0.551 | 0.549 | 0.763 | 0.561 | 0.573 | 0.739 |
| Left frontal operculum | 100.9 × SUVr -111.0 | 108.8 × SUVr -124.4 | 194.3 × SUVr -219.5 | 210.2 × SUVr -249.4 | 0.519 | 0.559 | 0.814 | 0.518 | 0.595 | 0.784 |
| Right frontal pole | 141.7 × SUVr -126.0 | 160.2 × SUVr -133.9 | 261.1 × SUVr -248.8 | 287.5 × SUVr -253.1 | 0.543 | 0.471 | 0.748 | 0.557 | 0.415 | 0.716 |
| Left frontal pole | 153.5 × SUVr -138.2 | 174.6 × SUVr -147.3 | 291.2 × SUVr -280.8 | 321.4 × SUVr -285.8 | 0.527 | 0.490 | 0.724 | 0.543 | 0.431 | 0.681 |
| Right fusiform gyrus | 121.2 × SUVr -132.8 | 133.8 × SUVr -148.4 | 220.8 × SUVr -242.3 | 235.5 × SUVr -264.7 | 0.549 | 0.496 | 0.826 | 0.568 | 0.494 | 0.818 |
| Left fusiform gyrus | 131.7 × SUVr -143.8 | 144.2 × SUVr -159.5 | 267.6 × SUVr -292.9 | 287.4 × SUVr -323.4 | 0.492 | 0.557 | 0.817 | 0.502 | 0.570 | 0.805 |
| Right gyrus rectus | 90.5 × SUVr -92.9 | 96.7 × SUVr -98.8 | 180.9 × SUVr -192.3 | 193.6 × SUVr -205.2 | 0.500 | 0.550 | 0.769 | 0.499 | 0.550 | 0.758 |
| Left gyrus rectus | 91.4 × SUVr -95.3 | 98.3 × SUVr -103.0 | 188.0 × SUVr -202.2 | 197.9 × SUVr -213.9 | 0.486 | 0.568 | 0.837 | 0.497 | 0.560 | 0.823 |
| Right inferior occipital gyrus | 123.0 × SUVr -135.1 | 137.2 × SUVr -150.5 | 226.7 × SUVr -254.7 | 245.9 × SUVr -278.5 | 0.543 | 0.527 | 0.858 | 0.558 | 0.520 | 0.845 |
| Left inferior occipital gyrus | 129.2 × SUVr -141.8 | 143.8 × SUVr -157.8 | 257.6 × SUVr -290.3 | 288.3 × SUVr -329.8 | 0.501 | 0.577 | 0.812 | 0.499 | 0.597 | 0.793 |
| Right inferior temporal gyrus | 98.6 × SUVr -103.6 | 108.5 × SUVr -113.6 | 174.8 × SUVr -191.5 | 190.9 × SUVr -210.8 | 0.564 | 0.503 | 0.881 | 0.568 | 0.509 | 0.868 |
| Left inferior temporal gyrus | 102.7 × SUVr -107.6 | 114.0 × SUVr -119.6 | 199.4 × SUVr -217.3 | 221.2 × SUVr -243.8 | 0.515 | 0.550 | 0.873 | 0.515 | 0.561 | 0.864 |

| | | | | | | | | | | |
|---|---|---|---|---|---|---|---|---|---|---|
| Right lingual gyrus | 259.2 × SUVr -298.2 | 282.7 × SUVr -329.7 | 409.6 × SUVr -446.0 | 441.8 × SUVr -495.0 | 0.633 | 0.361 | 0.820 | 0.640 | 0.374 | 0.817 |
| Left lingual gyrus | 243.7 × SUVr -274.6 | 262.5 × SUVr -299.4 | 408.6 × SUVr -443.6 | 425.2 × SUVr -471.9 | 0.596 | 0.414 | 0.701 | 0.617 | 0.406 | 0.704 |
| Right lateral orbital gyrus | 130.9 × SUVr -135.1 | 144.0 × SUVr -146.2 | 235.4 × SUVr -272.7 | 245.9 × SUVr -282.4 | 0.556 | 0.585 | 0.774 | 0.586 | 0.554 | 0.756 |
| Left lateral orbital gyrus | 136.3 × SUVr -141.5 | 150.6 × SUVr -156.8 | 264.9 × SUVr -295.8 | 282.8 × SUVr -321.8 | 0.515 | 0.583 | 0.816 | 0.533 | 0.584 | 0.809 |
| Right middle cingulate gyrus | 98.9 × SUVr -115.9 | 109.7 × SUVr -133.5 | 203.2 × SUVr -246.5 | 221.6 × SUVr -283.4 | 0.487 | 0.643 | 0.807 | 0.495 | 0.676 | 0.794 |
| Left middle cingulate gyrus | 103.9 × SUVr -125.3 | 115.7 × SUVr -144.1 | 211.7 × SUVr -259.0 | 231.5 × SUVr -298.8 | 0.491 | 0.631 | 0.828 | 0.500 | 0.668 | 0.812 |
| Right medial frontal cortex | 68.9 × SUVr -74.5 | 72.5 × SUVr -80.8 | 145.5 × SUVr -161.4 | 148.9 × SUVr -174.0 | 0.473 | 0.597 | 0.840 | 0.487 | 0.625 | 0.849 |
| Left medial frontal cortex | 70.2 × SUVr -76.2 | 74.7 × SUVr -84.0 | 150.5 × SUVr -169.2 | 156.1 × SUVr -183.9 | 0.466 | 0.618 | 0.844 | 0.479 | 0.640 | 0.847 |
| Right middle frontal gyrus | 90.7 × SUVr -96.6 | 100.7 × SUVr -107.5 | 165.3 × SUVr -191.0 | 182.7 × SUVr -212.6 | 0.548 | 0.571 | 0.869 | 0.551 | 0.575 | 0.860 |
| Left middle frontal gyrus | 91.2 × SUVr -95.3 | 101.2 × SUVr -106.4 | 166.4 × SUVr -186.3 | 183.9 × SUVr -208.6 | 0.548 | 0.547 | 0.907 | 0.550 | 0.556 | 0.901 |
| Right middle occipital gyrus | 110.3 × SUVr -116.9 | 123.2 × SUVr -130.1 | 199.5 × SUVr -224.3 | 215.6 × SUVr -241.7 | 0.553 | 0.538 | 0.880 | 0.571 | 0.518 | 0.867 |
| Left middle occipital gyrus | 111.6 × SUVr -117.3 | 124.5 × SUVr -130.8 | 225.2 × SUVr -253.7 | 246.3 × SUVr -278.9 | 0.496 | 0.606 | 0.857 | 0.505 | 0.601 | 0.829 |
| Right medial orbital gyrus | 115.5 × SUVr -128.4 | 126.1 × SUVr -141.7 | 229.4 × SUVr -267.1 | 243.6 × SUVr -289.3 | 0.503 | 0.605 | 0.782 | 0.518 | 0.606 | 0.772 |
| Left medial orbital gyrus | 109.4 × SUVr -118.9 | 119.6 × SUVr -130.4 | 214.8 × SUVr -254.1 | 229.9 × SUVr -274.3 | 0.509 | 0.629 | 0.818 | 0.520 | 0.626 | 0.806 |
| Right postcentral gyrus medial segment | 120.4 × SUVr -140.1 | 134.0 × SUVr -158.5 | 186.0 × SUVr -218.4 | 212.4 × SUVr -255.3 | 0.648 | 0.421 | 0.802 | 0.631 | 0.456 | 0.770 |
| Left postcentral gyrus medial segment | 136.1 × SUVr -162.5 | 148.7 × SUVr -176.6 | 254.8 × SUVr -304.8 | 287.7 × SUVr -342.3 | 0.534 | 0.559 | 0.716 | 0.517 | 0.576 | 0.676 |
| Right precentral gyrus medial segment | 135.6 × SUVr -161.7 | 150.8 × SUVr -185.2 | 248.7 × SUVr -306.1 | 283.4 × SUVr -365.3 | 0.545 | 0.581 | 0.770 | 0.532 | 0.636 | 0.734 |
| Left precentral gyrus medial segment | 144.1 × SUVr -177.0 | 160.9 × SUVr -202.3 | 249.2 × SUVr -306.7 | 279.5 × SUVr -356.8 | 0.578 | 0.520 | 0.757 | 0.576 | 0.553 | 0.738 |
| Right superior frontal gyrus medial segment | 77.4 × SUVr -85.5 | 83.8 × SUVr -94.8 | 155.7 × SUVr -175.2 | 168.7 × SUVr -197.8 | 0.497 | 0.576 | 0.848 | 0.497 | 0.610 | 0.830 |
| Left superior frontal gyrus medial segment | 84.6 × SUVr -94.3 | 92.6 × SUVr -105.4 | 180.9 × SUVr -203.5 | 195.6 × SUVr -226.1 | 0.468 | 0.604 | 0.843 | 0.473 | 0.617 | 0.825 |
| Right middle temporal gyrus | 93.2 × SUVr -96.7 | 103.6 × SUVr -109.2 | 173.2 × SUVr -191.1 | 191.2 × SUVr -215.7 | 0.538 | 0.545 | 0.894 | 0.542 | 0.557 | 0.888 |
| Left middle temporal gyrus | 96.1 × SUVr -98.8 | 106.7 × SUVr -111.5 | 190.8 × SUVr -207.8 | 211.8 × SUVr -235.5 | 0.504 | 0.571 | 0.859 | 0.504 | 0.586 | 0.854 |
| Right occipital pole | 238.9 × SUVr -247.7 | 275.5 × SUVr -270.8 | 375.7 × SUVr -388.7 | 418.0 × SUVr -412.0 | 0.636 | 0.375 | 0.789 | 0.659 | 0.338 | 0.728 |
| Left occipital pole | 228.6 × SUVr -229.3 | 265.2 × SUVr -252.6 | 455.5 × SUVr -481.5 | 509.4 × SUVr -511.4 | 0.502 | 0.554 | 0.773 | 0.521 | 0.508 | 0.731 |
| Right occipital fusiform gyrus | 168.0 × SUVr -200.0 | 182.6 × SUVr -220.7 | 284.5 × SUVr -325.2 | 307.0 × SUVr -368.1 | 0.590 | 0.440 | 0.705 | 0.595 | 0.480 | 0.714 |

| | | | | | | | | | | |
|---|---|---|---|---|---|---|---|---|---|---|
| Left occipital fusiform gyrus | 169.4 × SUVr -202.2 | 183.1 × SUVr -222.1 | 320.1 × SUVr -379.5 | 340.8 × SUVr -420.8 | 0.529 | 0.554 | 0.656 | 0.537 | 0.583 | 0.657 |
| Right opercular part of the inferior frontal gyrus | 112.7 × SUVr -123.1 | 125.6 × SUVr -139.7 | 194.1 × SUVr -222.4 | 214.4 × SUVr -251.6 | 0.581 | 0.512 | 0.859 | 0.586 | 0.522 | 0.843 |
| Left opercular part of the inferior frontal gyrus | 115.5 × SUVr -123.3 | 128.7 × SUVr -141.6 | 243.0 × SUVr -266.0 | 272.3 × SUVr -311.6 | 0.475 | 0.587 | 0.813 | 0.473 | 0.625 | 0.773 |
| Right orbital part of the inferior frontal gyrus | 111.8 × SUVr -121.8 | 122.3 × SUVr -136.1 | 195.1 × SUVr -223.6 | 210.2 × SUVr -247.8 | 0.573 | 0.522 | 0.763 | 0.582 | 0.531 | 0.749 |
| Left orbital part of the inferior frontal gyrus | 108.8 × SUVr -113.7 | 118.5 × SUVr -125.7 | 201.0 × SUVr -215.9 | 215.0 × SUVr -239.5 | 0.541 | 0.508 | 0.878 | 0.551 | 0.529 | 0.877 |
| Right posterior cingulate gyrus | 102.0 × SUVr -122.4 | 113.2 × SUVr -139.9 | 203.2 × SUVr -239.5 | 222.1 × SUVr -276.0 | 0.502 | 0.577 | 0.810 | 0.510 | 0.613 | 0.791 |
| Left posterior cingulate gyrus | 102.4 × SUVr -122.6 | 114.2 × SUVr -141.4 | 207.2 × SUVr -242.6 | 231.1 × SUVr -286.9 | 0.494 | 0.579 | 0.801 | 0.494 | 0.630 | 0.777 |
| Right precuneus | 84.5 × SUVr -95.4 | 90.7 × SUVr -104.9 | 167.0 × SUVr -187.2 | 176.9 × SUVr -205.9 | 0.506 | 0.550 | 0.864 | 0.513 | 0.571 | 0.857 |
| Left precuneus | 82.3 × SUVr -93.5 | 88.3 × SUVr -102.4 | 163.0 × SUVr -184.3 | 173.1 × SUVr -202.3 | 0.505 | 0.557 | 0.853 | 0.510 | 0.577 | 0.849 |
| Right parahippocampal gyrus | 282.3 × SUVr -319.8 | 323.1 × SUVr -365.6 | 562.3 × SUVr -586.6 | 657.1 × SUVr -694.6 | 0.502 | 0.474 | 0.675 | 0.492 | 0.501 | 0.633 |
| Left parahippocampal gyrus | 336.2 × SUVr -371.3 | 375.1 × SUVr -416.0 | 702.5 × SUVr -722.9 | 774.0 × SUVr -798.6 | 0.479 | 0.500 | 0.606 | 0.485 | 0.494 | 0.569 |
| Right posterior insula | 146.0 × SUVr -167.5 | 160.0 × SUVr -188.5 | 247.6 × SUVr -273.6 | 271.9 × SUVr -309.6 | 0.590 | 0.429 | 0.790 | 0.588 | 0.445 | 0.765 |
| Left posterior insula | 139.3 × SUVr -153.3 | 150.4 × SUVr -170.7 | 275.4 × SUVr -291.1 | 291.4 × SUVr -321.1 | 0.506 | 0.500 | 0.759 | 0.516 | 0.516 | 0.749 |
| Right parietal operculum | 113.4 × SUVr -132.1 | 125.0 × SUVr -151.7 | 192.4 × SUVr -238.5 | 212.5 × SUVr -280.0 | 0.589 | 0.553 | 0.803 | 0.588 | 0.604 | 0.783 |
| Left parietal operculum | 113.5 × SUVr -129.1 | 124.9 × SUVr -148.6 | 206.6 × SUVr -247.1 | 230.9 × SUVr -295.0 | 0.549 | 0.571 | 0.796 | 0.541 | 0.634 | 0.762 |
| Right postcentral gyrus | 191.5 × SUVr -214.5 | 216.3 × SUVr -240.4 | 312.2 × SUVr -367.9 | 346.9 × SUVr -406.3 | 0.614 | 0.491 | 0.833 | 0.624 | 0.478 | 0.808 |
| Left postcentral gyrus | 187.4 × SUVr -203.3 | 210.0 × SUVr -227.4 | 332.8 × SUVr -384.2 | 370.4 × SUVr -427.6 | 0.563 | 0.544 | 0.795 | 0.567 | 0.540 | 0.777 |
| Right posterior orbital gyrus | 125.6 × SUVr -135.4 | 133.5 × SUVr -144.6 | 218.2 × SUVr -231.9 | 227.2 × SUVr -244.8 | 0.576 | 0.442 | 0.852 | 0.588 | 0.441 | 0.853 |
| Left posterior orbital gyrus | 119.1 × SUVr -126.1 | 127.1 × SUVr -136.2 | 218.7 × SUVr -230.3 | 233.2 × SUVr -251.3 | 0.544 | 0.477 | 0.820 | 0.545 | 0.494 | 0.810 |
| Right planum polare | 141.8 × SUVr -157.2 | 149.8 × SUVr -172.1 | 259.2 × SUVr -272.3 | 283.2 × SUVr -315.9 | 0.547 | 0.444 | 0.706 | 0.529 | 0.508 | 0.669 |
| Left planum polare | 147.2 × SUVr -159.0 | 158.2 × SUVr -178.7 | 298.2 × SUVr -299.1 | 310.7 × SUVr -336.7 | 0.493 | 0.470 | 0.663 | 0.509 | 0.508 | 0.668 |
| Right precentral gyrus | 203.5 × SUVr -233.2 | 229.9 × SUVr -263.4 | 323.4 × SUVr -394.0 | 375.0 × SUVr -458.3 | 0.629 | 0.497 | 0.781 | 0.613 | 0.520 | 0.735 |
| Left precentral gyrus | 210.7 × SUVr -236.1 | 238.2 × SUVr -269.6 | 378.6 × SUVr -453.4 | 431.2 × SUVr -523.1 | 0.556 | 0.574 | 0.763 | 0.552 | 0.588 | 0.729 |
| Right planum temporale | 116.0 × SUVr -128.0 | 123.5 × SUVr -139.7 | 230.9 × SUVr -259.6 | 246.3 × SUVr -288.3 | 0.502 | 0.570 | 0.763 | 0.501 | 0.604 | 0.720 |

| | | | | | | | | | | |
|---|---|---|---|---|---|---|---|---|---|---|
| Left planum temporale | 113.7 × SUVr -123.8 | 121.9 × SUVr -137.6 | 222.3 × SUVr -246.5 | 230.8 × SUVr -266.3 | 0.511 | 0.552 | 0.817 | 0.528 | 0.557 | 0.807 |
| Right subcallosal area | 112.6 × SUVr -116.7 | 117.5 × SUVr -121.7 | 240.7 × SUVr -238.8 | 249.9 × SUVr -250.1 | 0.468 | 0.507 | 0.725 | 0.470 | 0.514 | 0.718 |
| Left subcallosal area | 114.9 × SUVr -120.3 | 121.1 × SUVr -127.2 | 245.5 × SUVr -247.7 | 255.6 × SUVr -258.5 | 0.468 | 0.519 | 0.730 | 0.474 | 0.514 | 0.730 |
| Right superior frontal gyrus | 96.9 × SUVr -101.6 | 109.5 × SUVr -114.2 | 177.5 × SUVr -197.7 | 204.1 × SUVr -226.4 | 0.546 | 0.541 | 0.822 | 0.537 | 0.550 | 0.807 |
| Left superior frontal gyrus | 100.2 × SUVr -105.1 | 113.7 × SUVr -119.3 | 183.5 × SUVr -204.3 | 210.6 × SUVr -235.2 | 0.546 | 0.540 | 0.874 | 0.540 | 0.550 | 0.855 |
| Right supplementary motor cortex | 110.0 × SUVr -125.5 | 121.7 × SUVr -142.5 | 215.1 × SUVr -249.3 | 246.8 × SUVr -297.5 | 0.511 | 0.576 | 0.843 | 0.493 | 0.628 | 0.803 |
| Left supplementary motor cortex | 116.8 × SUVr -136.8 | 128.0 × SUVr -152.5 | 223.9 × SUVr -262.6 | 244.1 × SUVr -293.9 | 0.522 | 0.562 | 0.828 | 0.525 | 0.579 | 0.799 |
| Right supramarginal gyrus | 110.5 × SUVr -115.7 | 123.9 × SUVr -130.2 | 202.4 × SUVr -226.1 | 224.4 × SUVr -251.6 | 0.546 | 0.546 | 0.855 | 0.552 | 0.541 | 0.841 |
| Left supramarginal gyrus | 112.5 × SUVr -115.2 | 125.3 × SUVr -129.6 | 227.2 × SUVr -249.0 | 250.7 × SUVr -277.2 | 0.495 | 0.589 | 0.849 | 0.500 | 0.589 | 0.840 |
| Right superior occipital gyrus | 244.7 × SUVr -271.3 | 279.1 × SUVr -312.5 | 427.2 × SUVr -500.9 | 483.0 × SUVr -579.9 | 0.573 | 0.537 | 0.742 | 0.578 | 0.553 | 0.711 |
| Left superior occipital gyrus | 238.9 × SUVr -267.6 | 270.7 × SUVr -305.1 | 412.2 × SUVr -478.9 | 454.4 × SUVr -537.8 | 0.580 | 0.513 | 0.736 | 0.596 | 0.512 | 0.703 |
| Right superior parietal lobule | 104.7 × SUVr -115.2 | 115.3 × SUVr -127.3 | 195.1 × SUVr -222.9 | 216.2 × SUVr -248.4 | 0.537 | 0.552 | 0.855 | 0.533 | 0.560 | 0.840 |
| Left superior parietal lobule | 101.4 × SUVr -111.5 | 112.4 × SUVr -124.6 | 198.8 × SUVr -229.6 | 221.8 × SUVr -258.7 | 0.510 | 0.594 | 0.853 | 0.507 | 0.605 | 0.839 |
| Right superior temporal gyrus | 97.1 × SUVr -101.2 | 108.1 × SUVr -114.0 | 183.0 × SUVr -204.7 | 200.6 × SUVr -228.2 | 0.530 | 0.565 | 0.848 | 0.539 | 0.569 | 0.837 |
| Left superior temporal gyrus | 100.2 × SUVr -105.1 | 111.7 × SUVr -118.3 | 193.3 × SUVr -214.2 | 215.1 × SUVr -242.6 | 0.518 | 0.565 | 0.841 | 0.519 | 0.578 | 0.836 |
| Right temporal pole | 160.4 × SUVr -155.1 | 176.5 × SUVr -169.5 | 296.3 × SUVr -288.4 | 315.2 × SUVr -304.7 | 0.541 | 0.450 | 0.766 | 0.560 | 0.429 | 0.757 |
| Left temporal pole | 170.2 × SUVr -164.1 | 188.2 × SUVr -180.3 | 331.0 × SUVr -322.7 | 359.2 × SUVr -347.9 | 0.514 | 0.479 | 0.807 | 0.524 | 0.467 | 0.788 |
| Right triangular part of the inferior frontal gyrus | 107.0 × SUVr -113.7 | 119.0 × SUVr -126.3 | 189.5 × SUVr -218.1 | 206.0 × SUVr -239.3 | 0.565 | 0.551 | 0.835 | 0.578 | 0.548 | 0.814 |
| Left triangular part of the inferior frontal gyrus | 102.5 × SUVr -104.5 | 114.5 × SUVr -118.9 | 190.5 × SUVr -207.3 | 211.4 × SUVr -235.8 | 0.538 | 0.540 | 0.895 | 0.542 | 0.553 | 0.876 |
| Right transverse temporal gyrus | 141.0 × SUVr -164.2 | 147.7 × SUVr -173.1 | 258.7 × SUVr -302.6 | 271.6 × SUVr -319.2 | 0.545 | 0.535 | 0.783 | 0.544 | 0.538 | 0.784 |
| Left transverse temporal gyrus | 131.7 × SUVr -149.2 | 138.0 × SUVr -158.4 | 252.0 × SUVr -281.6 | 261.1 × SUVr -296.7 | 0.522 | 0.525 | 0.772 | 0.529 | 0.530 | 0.761 |

Regional target regions represent predefined MUSE gray matter regions or composite regions. CL, Centiloid; MUSE, MUlti-atlas region Segmentation utilizing Ensembles; PiB, [$^{11}$C]Pittsburgh compound B; AV-45, [$^{18}$F]florbetapir; PLS-PVC, parallel level set regularization-based partial-volume correction; NoPVC, without partial-volume correction; $R^2$, coefficient of determination.

#### Supplementary Table 14. Diagnostic group separation of MUSE-derived regional Centiloid measurements in the independent application cohorts

| Cohort/Tracer | Region | AD N | CN N | NoPVC AD CL, mean ± SD | NoPVC CN CL, mean ± SD | NoPVC Hedges *g* | NoPVC *q* | PLS-PVC AD CL, mean ± SD | PLS-PVC CN CL, mean ± SD | PLS-PVC Hedges *g* | PLS-PVC *q* | Δ*g* |
|---|---|---|---|---|---|---|---|---|---|---|---|---|
| Knight-ADRC/PiB | Posterior cingulate | 18 | 15 | 104.9 ± 77.2 | 6.9 ± 31.4 | 1.569 | <0.001 | 107.9 ± 78.7 | 5.8 ± 27.8 | 1.627 | <0.001 | 0.057 |
| | Lateral temporal GM | 18 | 15 | 94.2 ± 70.3 | −4.2 ± 15.0 | 1.811 | <0.001 | 100.2 ± 74.7 | −4.4 ± 13.7 | 1.819 | <0.001 | 0.008 |
| | Frontal GM | 18 | 15 | 93.4 ± 67.5 | −8.0 ± 15.1 | 1.939 | <0.001 | 100.2 ± 72.9 | −6.3 ± 14.1 | 1.897 | <0.001 | −0.042 |
| | Precuneus | 18 | 15 | 99.7 ± 60.4 | 3.9 ± 19.6 | 2.006 | <0.001 | 104.8 ± 65.2 | 3.8 ± 19.9 | 1.967 | <0.001 | −0.039 |
| | Parietal GM | 18 | 15 | 98.2 ± 65.4 | 0.9 ± 15.2 | 1.920 | <0.001 | 103.8 ± 70.9 | 1.9 ± 14.9 | 1.860 | <0.001 | −0.060 |
| | Occipital GM | 18 | 15 | 95.2 ± 81.6 | 5.5 ± 25.1 | 1.395 | <0.001 | 98.9 ± 85.9 | 4.0 ± 23.2 | 1.412 | <0.001 | 0.018 |
| ADNI/AV-45 | Posterior cingulate | 119 | 119 | 35.8 ± 111.8 | −10.4 ± 71.8 | 0.490 | <0.001 | 44.2 ± 103.0 | −5.5 ± 63.2 | 0.580 | <0.001 | 0.091 |
| | Lateral temporal GM | 119 | 119 | 62.1 ± 78.4 | 5.3 ± 56.8 | 0.827 | <0.001 | 65.9 ± 77.1 | 7.0 ± 54.3 | 0.882 | <0.001 | 0.055 |
| | Frontal GM | 119 | 119 | 60.9 ± 72.9 | 4.0 ± 49.7 | 0.909 | <0.001 | 66.4 ± 72.2 | 7.6 ± 47.4 | 0.960 | <0.001 | 0.052 |
| | Precuneus | 119 | 119 | 59.7 ± 82.6 | 3.1 ± 53.9 | 0.809 | <0.001 | 64.4 ± 80.8 | 6.1 ± 51.3 | 0.859 | <0.001 | 0.050 |
| | Parietal GM | 119 | 119 | 61.9 ± 86.5 | 3.9 ± 59.6 | 0.779 | <0.001 | 67.2 ± 85.3 | 7.7 ± 56.9 | 0.817 | <0.001 | 0.039 |
| | Occipital GM | 119 | 119 | 103.6 ± 82.7 | 27.6 ± 62.2 | 1.036 | <0.001 | 106.7 ± 81.8 | 28.8 ± 60.8 | 1.077 | <0.001 | 0.041 |

CL values are presented as mean ± SD. q values correspond to Welch-test results adjusted for multiple comparisons across the six predefined targets. Knight-ADRC PiB, 30-min acquisitions (18 AD, 15 CN); ADNI AV-45, matched cohort (119 AD, 119 CN). Δg was calculated as PLS-PVC minus NoPVC. AV-45, [$^{18}$F]florbetapir; CL, Centiloid; GM, gray matter; PiB, [$^{11}$C]Pittsburgh compound B; PLS-PVC, parallel level set regularization-based partial-volume correction; NoPVC, without partial-volume correction.

**Supplementary Table 15. Clinical associations of MUSE-derived regional Centiloid measurements in the independent application cohorts**

| Cohort/Tracer | Region | CDR-SB | | | | MMSE | | | |
|---|---|---|---|---|---|---|---|---|---|
| | | **NoPVC adjusted ρ** | **NoPVC adjusted *q*** | **PLS-PVC adjusted *ρ*** | **PLS-PVC adjusted *q*** | **NoPVC adjusted *ρ*** | **NoPVC adjusted *q*** | **PLS-PVC adjusted *ρ*** | **PLS-PVC adjusted *q*** |
| Knight-ADRC/PiB | Posterior cingulate | 0.468 | 1.10E-02 | 0.448 | 1.57E-02 | -0.451 | 3.61E-02 | -0.440 | 3.84E-02 |
| | Lateral temporal GM | 0.555 | 4.36E-03 | 0.560 | 3.98E-03 | -0.392 | 4.23E-02 | -0.389 | 4.47E-02 |
| | Frontal GM | 0.576 | 4.36E-03 | 0.559 | 3.98E-03 | -0.418 | 3.61E-02 | -0.404 | 4.47E-02 |
| | Precuneus | 0.438 | 1.54E-02 | 0.435 | 1.63E-02 | -0.439 | 3.61E-02 | -0.437 | 3.84E-02 |
| | Parietal GM | 0.495 | 9.38E-03 | 0.498 | 9.97E-03 | -0.433 | 3.61E-02 | -0.432 | 3.84E-02 |
| | Occipital GM | 0.488 | 9.38E-03 | 0.485 | 9.97E-03 | -0.268 | 1.59E-01 | -0.256 | 1.80E-01 |
| ADNI/AV-45 | Posterior cingulate | 0.243 | 1.67E-04 | 0.262 | 4.93E-05 | -0.266 | 3.66E-05 | -0.280 | 1.27E-05 |
| | Lateral temporal GM | 0.315 | 1.29E-06 | 0.337 | 2.47E-07 | -0.358 | 3.30E-08 | -0.376 | 5.02E-09 |
| | Frontal GM | 0.342 | 2.21E-07 | 0.361 | 3.75E-08 | -0.367 | 2.07E-08 | -0.388 | 2.25E-09 |
| | Precuneus | 0.317 | 1.29E-06 | 0.331 | 3.10E-07 | -0.340 | 1.38E-07 | -0.350 | 5.11E-08 |
| | Parietal GM | 0.301 | 3.05E-06 | 0.307 | 1.99E-06 | -0.330 | 2.61E-07 | -0.339 | 1.15E-07 |
| | Occipital GM | 0.434 | 1.87E-11 | 0.453 | 1.57E-12 | -0.453 | 1.58E-12 | -0.473 | 1.04E-13 |

Values are partial Spearman correlations adjusted for age, sex, and education; *q* values represent multiple-comparison-adjusted significance values. CDR-SB, Clinical Dementia Rating Sum of Boxes; MMSE, Mini-Mental State Examination; other abbreviations as defined above; NoPVC, without partial-volume correction.
